# Dense sampling of choices links high learning rates to obesity and low reward sensitivity to binge eating

**DOI:** 10.1101/2025.06.19.25329936

**Authors:** Anne Kühnel, Juliane Zietz, Johanna Theuer, Monja P. Neuser, Vanessa Teckentrup, Franziska K. Müller, Peter Dayan, Jennifer Svaldi, Nils B. Kroemer

## Abstract

Mounting evidence shows that obesity is associated with alterations in dopamine transmission. However, in humans, corresponding changes in dopamine-dependent behavior, such as reward learning with increasing BMI, have not been conclusively established and dissociated from pathological eating behavior. Here, we provide a principled assessment of differences in reinforcement learning (RL) related to obesity or pathological eating, such as binge eating (BE), using a large sample of 370 participants enriched for high BMI and BE. To dissociate trait and state components of RL, participants completed up to 31 runs of a novel RL game across weeks (1,486,950 observed choices). We used hierarchical Bayesian models to estimate the mean and variability of parameters for each individual. Across runs, higher BMI was predicted by higher and more variable learning rates using elastic net models (*ΔR*^2^=.19; *p_perm_*<.0001) although performance did not differ (*r*=-0.04, *p*_boot_=.46). In contrast, BE and the dimension loss of control were characterized by lower and more variable reward sensitivity (*ΔR*^2^=.19, controlled for BMI; *p_perm_*<.0001) leading to fewer points (*r*=.10, *p*_boot_=.030). Crucially, patients with BE disorder also showed attenuated reward sensitivity in the nucleus accumbens during anticipation in an effort task that required no learning (*p*=.006). We conclude that obesity-related differences in reward learning are best explained by changes in learning, whereas BE and loss of control were mostly driven by reduced reward sensitivity. Notably, repeated assessments revealed that increased variability across states contribute to both obesity and BE. Our findings highlight the necessity to complement neurobiological research with behavioral precision mapping to derive mechanistic insight into multidimensional disorders such as obesity and BE disorder.

## Introduction

Obesity is one of the main preventable causes of death in developed countries today ^1,2^. To maintain energy homeostasis, any foraging animal must learn over time which prospective sources of food are good and which sources are bad and how much food must be consumed to recuperate metabolized energy ^3–6^. However, in contrast to most foraging species, for us in today’s affluent societies, the omnipresence of rewards competing for our attention in today’s affluent societies poses a very different challenge for maintaining energy balance ^7^. Nevertheless, we still have to rely upon the same basic neurobiological mechanisms of reward learning, including aspects of the action of the neuromodulator dopamine e.g., ^8,9–15^. Crucially, there is conclusive evidence from both animal ^16–20^ and human research ^21–26^ showing alterations in dopaminergic neurotransmission in obesity, suggesting the possibility of a malign feedback loop. These conceptual links raise the question to what extent inter-individual differences in reward learning contribute to pathological eating behaviors such as binge eating (BE) and obesity ^23^.

Although the neurobiological link between energy metabolism, dopamine transmission, and obesity is well established, the reported evidence on associations between obesity and reward learning is so far mixed. Most studies have reported an impairment in learning for food ^27–30^ and monetary rewards ^27,31^. In some studies, these deficits in learning were more pronounced for losses ^27,31^, negative prediction errors ^32^, or specific to model-based control ^33^. Several other studies have reported no difference in average reward outcome signals ^34^ or probabilistic learning for food ^35^ or monetary rewards ^28^. Other studies have even reported enhanced learning for food ^36^ and monetary rewards ^36,37^. In addition to learning rates, decision-making in RL tasks is governed by the reward sensitivity that scales whether participants typically choose the option they value more ^38^. Perhaps surprisingly, reward sensitivity has been shown to be reduced for non-food rewards in obesity, leading to noisier choices ^32,39,40^, although self-report questionnaires of reward sensitivity show increased or similar levels ^41^. Taken together, the associations of BMI with altered learning behavior vary substantially across studies, raising the question whether any reliable pattern characterizes obesity and comorbid pathological eating behavior.

Taking a cue from other transdiagnostic approaches, dimensions of pathological eating behavior may better map onto specific learning and decision-making patterns than obesity, similar to other transdiagnostic dimensions ^42^. Loss of control over eating and BE are promising candidates to explain discrepancies across studies since they characterize a subset of patients with obesity. Patients with BE have been shown to perform worse in reversal learning tasks ^43^ and to rely more on model-free learning ^44,45^. Commonly, BE is often related to higher reward sensitivities ^46^, but most evidence comes from studies using passive viewing of rewards without instrumental behavior. The few studies investigating reward sensitivity with behavioral tasks in BE have mostly shown small effects, predominantly pointing to reduced reward sensitivity ^47–49^, although effects are sometimes valence-dependent e.g., win vs. loss, ^39^. Most of these studies only include either a normal weight or an obese control group, making it difficult to disentangle the effect of excessive weight from BE, although distinct associations have been reported ^39^. To conclude, dimensions of eating behavior including BE might have discernible signatures of reward learning compared to obesity, but they are rarely investigated with computational or predictive modeling techniques.

Although the association between obesity as well as eating behavior and reward learning is assumed, most evidence for obesity- or eating-behavior-related changes in reward learning comes from comparatively small studies (*N*s 30-135) that rely on a limited number of runs to estimate latent parameters reflecting reward learning. However, previous research has indicated only moderate test-retest reliability of common RL parameters ^50^, suggesting that multiple runs may be necessary to better isolate trait and state components of reward learning from measurement noise ^51,52^. Moreover, relying on averages might miss relevant information as BE might be characterized by the variability of behavior over time with BE episodes occurring in extreme states ^53^. Again, this variability across states can only be quantified across multiple runs ^53^. Here, we close this gap by conducting an extensive assessment of reward learning over weeks in a large sample enriched for high BMI (i.e., a range of almost 50 units) and key symptoms of pathological eating, such as BE and excessive restraint, that were comprehensively assessed with questionnaires and EMA. To separate trait and state components of reward learning, we used a hierarchical Bayesian sampling technique to estimate average parameters as well as their dispersion over runs. Based on previous studies, we hypothesized that BMI would be negatively associated with reward learning, particularly within the range from normal weight to class-I obesity. In contrast, we expected that BE would be associated with lower and more variable reward sensitivity over time.

## Methods

### Participants

For the reported analyses, we used the data of 370 participants (299 women, *M*_age_ = 34.8 years ± 14.1, [18 – 68]; *M*_BMI_= 26.3 kg/m^2^ ± 7.1; [13.8 – 58.5]) from a study on trait and state markers of binge eating ^53^. Participants were recruited through the University of Tübingen’s email lists, online ads, and press releases. Since BE is more prevalent in women than to men, we oversampled women with pathological eating behavior to improve the estimation of associations with symptoms that would otherwise only occur in 5%-15% of a population sample ^54,55^. We screened all participants for their physical and mental health via phone interviews. All included participants completed at least 10 runs of the gamified RL task ^51^ and a set of questionnaires related to their eating behavior and mental health. Participants provided written informed consent before completing the set of questionnaires. They received either money (20€) or partial course credit in return for their participation in this part of the experiment. In addition to the fixed compensation, eligible participants were invited for further behavioral and neuroimaging visits as part of the larger study, where they would receive additional remuneration. The institutional review board of the Faculty of Medicine (i.e., the ethics committee), University of Tübingen, approved this study (reference number 393/2017BO2), which was conducted in accordance with the Declaration of Helsinki by the World Medical Association. The study protocol was preregistered at clinicaltrials.gov (NCT04184856). The analyses using reinforcement learning were not preregistered and are based on previous work establishing the model and hypotheses regarding variability ^51,53^.

We estimated a priori that we need to enroll at least 230 participants in this part of the study to achieve a high power 1-*β* = .87 to study small associations with learning parameters (effect size *r*=.2). Since more participants (*N*=370) completed the online assessment until the data freeze by the end of the project (2021/07/01), the power to investigate small effects was even higher (1-*β* = .98). Crucially, RL simulations of virtual agents suggested that our extensive repeated-measures design is highly sensitive to recover such latent differences due to the increased accuracy of the point estimates compared to conventional single-run designs.

### Gamified reinforcement learning task

To measure reliable trait components of reward learning, we developed a gamified RL task called “Influenca” ^51^. The task is similar to a classic RL task ^56^. Briefly, in each level (150 trials), participants had to find the best of two medications for a new virus to save as many lives as possible. In each trial, participants chose between the two medications. Each option was associated with a random reward magnitude, *f* ∈ [0, 100], independent of the win probability of each option, corresponding to the potential number of people cured. In the case of a win, the corresponding points were added to the total score or subtracted in case of a loss. The win probabilities of both options added up to one. In the first trial, one randomly chosen option was initialized with a win probability, *p*_A_ = .80, while the other option had the corresponding win probability of *p*_B_ = .20. After initialization, the win probabilities fluctuated as determined by a Gaussian random walk to prevent participants from learning an underlying task structure over 31 levels of the game.

To capture state effects, we complemented the task with a set of items to be answered prior to every run. The set of state items covered metabolic ratings (hunger, satiety, thirst), mood, and potential distractors as facets (e.g., stress, alertness, distraction by environment or thoughts). Moreover, participants were asked whether they had experienced a BE episode since they last played the game. We coded the app in the open language HAXE (Haxe Foundation, 2018), and the source code is available via a GitHub repository (https://github.com/VTeckentrup/mind-mosaic). We compiled game installers for different operating systems (i.e., Windows, Android, MacOS, Linux) to allow users to use their preferred device. These installers are hosted on our lab’s homepage (https://neuromadlab.com/de/influenca/) and provide a parallel version implemented in Pavlovia (https://github.com/neuromadlab/Tasks/tree/master/Influenca/Stable_version).

### Food questionnaires

To capture the inherent complexity of human eating behavior, we used 133 items belonging to 13 subscales from 6 commonly used questionnaires.: The Three-Factor Eating Questionnaire, TFEQ ^57^, the Power of Food Scale, PFS ^58^, the Yale Food Addiction Scale, YFAS ^59^, the Salzburg Emotional Eating Scale, SEES ^60^, the Salzburg Stress Eating Scale, SSES ^61^, and the Food Cravings Questionnaire – Trait, FCQ-T-r ^62^.

### Experimental procedure: online assessment

Participants were asked to install the app on their preferred personal devices. At registration, they had to enter an email address. Before launching the first run, participants were provided with detailed instructions of the game including the cover story. During the experimental phase, they could return to these instructions at any time via the game’s menu. Participants were instructed to play at different times of the day and in different (metabolic) states. To facilitate the sampling of reward learning over time, participants had to wait for at least 2 h after completing a run. After participants had completed 10 runs of the game, they were asked to answer a comprehensive set of online questionnaires to obtain an activation code that would unlock access to levels 11 to 31. Data of each run was stored locally on the participant’s device and synchronized (once the device had access to the Internet) with a database located at a server at the Department of Psychiatry and Psychotherapy in Tübingen.

To assess the physical and mental health of participants, we invited all players to phone screenings. This screening included questions from the Eating Disorder Examination, EDE ^64,65^, which was used to assess symptoms of pathological eating. Based on their eligibility for further lab-based assessments, participants were then invited for future visits.

### Data analysis

#### Computational modeling

Based on previous model comparisons ^51^, we used a reinforcement learning model including 4 parameters, two learning rates, reward sensitivity, and an additive scaling of points at stake ^66^. The learning rate, *α*, characterizes the speed of adaptation, describing how quickly participants update their choice preferences. This model (eq. 1a-c) uses two learning rates: One for trials in which the participant won points (*α_rew_*), and one for trials in which they lost points (*α_pun_*). The reward sensitivity parameter, *β*, describes the predictability of their choices or, conversely, the noisiness of their choices. High values of this parameter correspond to deterministic choices. Additionally, the model incorporates a mixture parameter, *λ*, which describes the relative importance of the potential reward (or loss) magnitude compared to the estimated win probability. Larger values of *λ* correspond to a higher influence of the estimated probability rather than the number of points at stake.

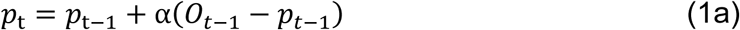

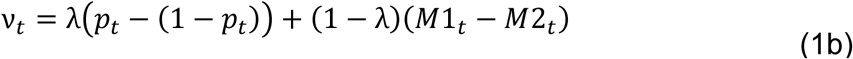

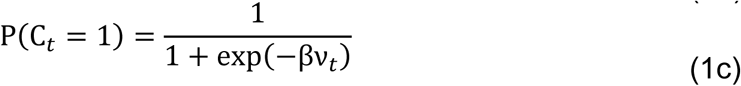

In eq. 1 ^66^, *p* refers to the estimated probability, *O* to the binary outcome of a trial (whether points were won or lost), *M1* and *M2* to the magnitude of each choice (points to be won or lost), and C to the participant’s choice, at trial *t*.

### Hierarchical Bayesian modeling

To fit person-level and run-level parameters to the data, we used hierarchical Bayesian models, since they provide regularization of the parameters at the level of the group and person. Briefly, in Bayesian estimation, posterior distributions of parameter estimates are approximated by sampling with Markov Chain Monte Carlo methods and the No-U-Turn Sampler (NUTS) ^67^. To capture the hierarchical structure of our data (i.e., trials nested within runs, runs within persons, and persons within groups), we built a model with three hierarchy levels (run, participant, group) based on the four-parameter additive model (eq. 1a-c) with two learning rates. The model used run-level priors for the four main model parameters (*α_win_*, *α_loss_*, *β*, *λ*) that are drawn from higher-level priors on the participant level (Fig. 1). In turn, they are then drawn from group-level hyperpriors. To define the prior distributions, we mostly used Beta- and Gamma-distributed priors (for full list of distributions see SI) as recommended ^68,69^. Since both the learning rates *α* and the mixture parameter *λ* vary between 0 and 1, priors were drawn from the Beta distribution (one for each participant). Each participant-level Beta prior distribution was parameterized by two hyperparameters, a1 and a2 with Beta(1+a1,1+a2). Both hyperparameters a1 and a2 were incremented by 1 to enforce a minimum value of the hyperparameters of 1 that ensures that the Beta distribution follows an inverted U-shape. The group-level hyperpriors used as hyperparameters of these Beta-distributed priors were selected from the Gamma distribution, which constrained them to be positive (i.e., smaller values would be preferred). Since the reward sensitivity has to be positive as well, the parameter *β* was also given Gamma-distributed priors with hyperpriors from the Gamma and Normal (transformed) distributions (for a list of all prior distributions, see SI). In the final model, to help with computational efficiency, *β* was also constrained so that it could not be higher than 15 (using Stan’s built-in constraint transforms) because higher values of the reward sensitivity would not make any appreciable difference ^51^. Models were implemented using Stan,^70^ through its interface to R, *rstan* ^71^. For these fits, all parameters were initialized to 0, and the model was fit with four chains with 4000 iterations each, with 1500 of those serving as warmup. To ensure a successful model fit, it was required that all chains converged with no divergences, *R̂* ≤ 1.05 (Gelman & Rubin, 1992), and the effective sampling size > 400 ^72^ for all parameters. In addition to these criteria and computational checks using diagnostic plots, we ensured that the model explained the data well by testing parameter recovery (Fig. S2) on simulated data with known parameters ^68,73^.

**Figure 1:**
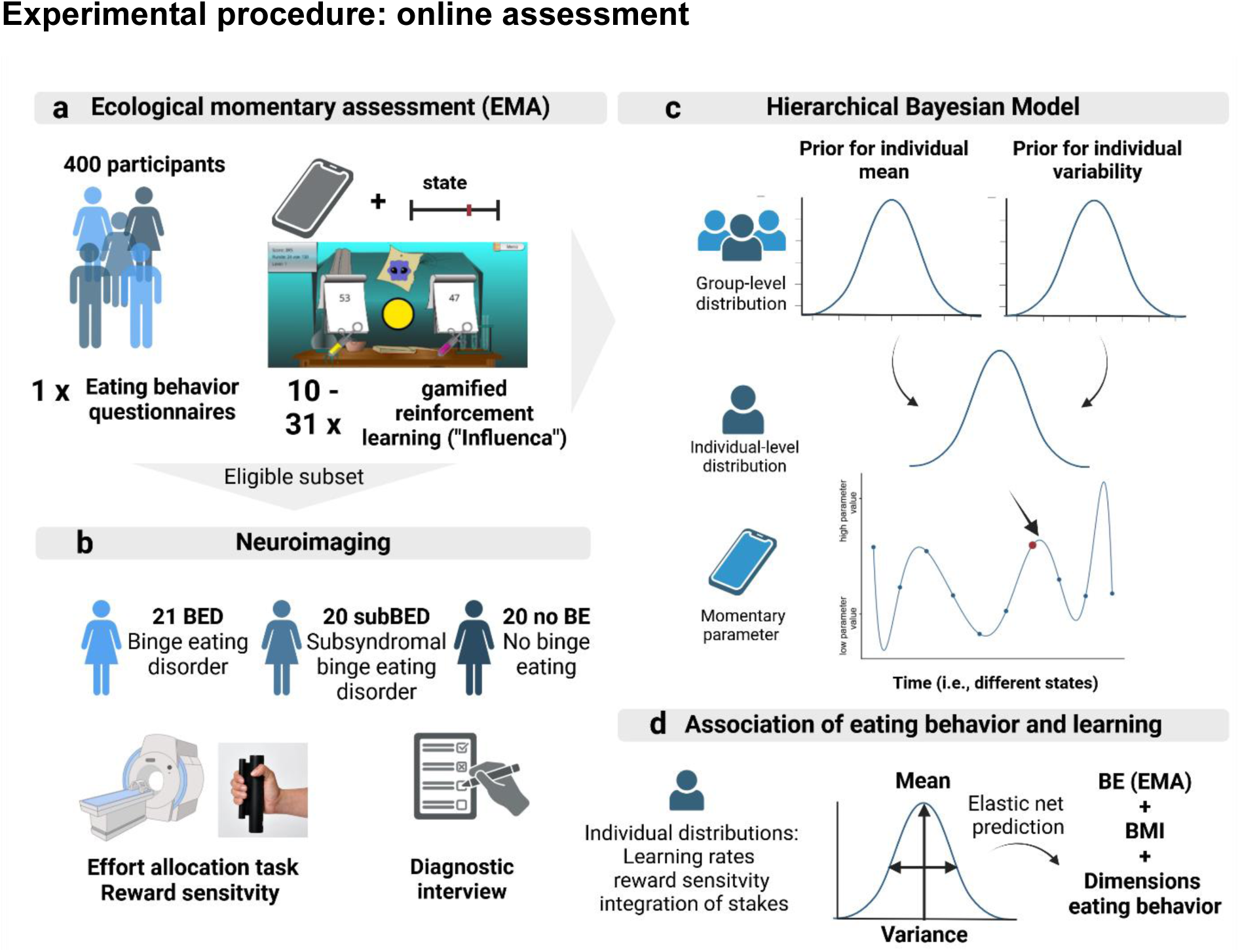
To determine distinct learning and decision-making profiles in obesity and binge eating (BE), we combined ecological momentary assessment (EMA) in a large online sample with a neuroimaging study. a: 400 participants completed a longitudinal online assessment including EMA questions on mood and metabolic states and a reinforcement learning task “Influenca” ^51^. Each participant completed at least 10 runs of the game. b: We invited a subsample of *n*=61 participants to a neuroimaging visit to measure reward response in the nucleus accumbens in an effort allocation task ^63^. c: We used hierarchical Bayesian modeling to explain choice behavior based on a reinforcement learning model. Crucially, the model included individual distributions for each parameter that allowed participants to differ in their average (i.e., mean) behavior as well as their variability (i.e., variance) across runs. The distributions depicted here are for illustration only and do not represent the chosen prior distributions. d: Last, we used multivariate elastic net models to determine which features of individual means and variances in learning parameters explained self-reported binge eating (BE EMA) or the self-reported body mass index (BMI). Figure created with BioRender.com.

#### Trait- and state characteristics of reinforcement learning

To quantify trait-like average learning behavior and its variability, the participant-level first (i.e., mean) and second (i.e., standard deviation) moment are directly sampled as transformed variables in the STAN model with the following formulas using the STAN parametrization of the distributions. For the participant-level distributions of the learning rates and lambda parameter (constraint between 0 and 1) the mean and standard deviation are calculated by:

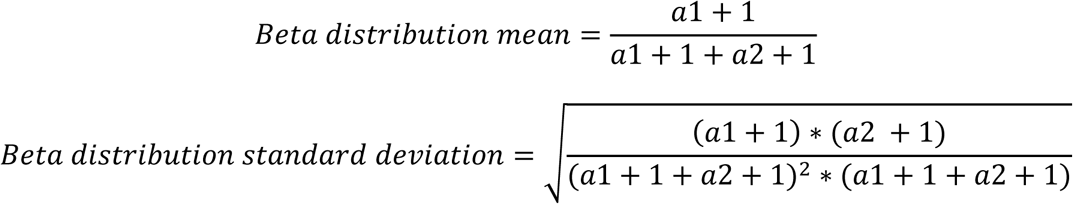

The participant-level mean and standard deviation for the reward sensitivity, β, are derived from the Gamma distribution as follows:

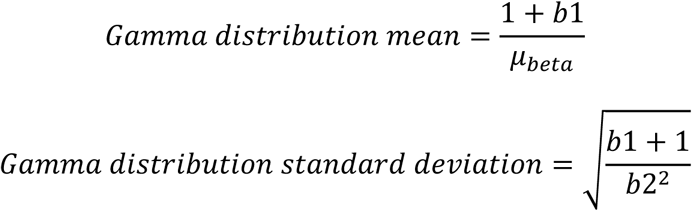

Here, a1 and a2 refer to the α and β parameters of a Beta distribution, and b1 and b2 are the shape and rate parameters of a Gamma distribution, respectively. Since the chosen prior distributions and their parametrization led to substantial correlations between estimates of the mean and their variance, we residualized the variance estimates by the mean for all further analyses.

To quantify state effects on learning parameters (e.g., of the current self-reported states or improvements over runs), we estimated mixed-effects models based on the average run-level posterior estimates of the hierarchical Bayesian model. The linear mixed-effects models were estimated using Bayesian sampling in *brms* ^74^. To capture meta-learning across runs, we included the intercept and either a log-transformed (learning rates and reward sensitivity) or exponentially transformed (*λ*, additive scaling) run counter as random effects (varying by ID) to the model. In addition, the variance estimate was also allowed to vary by participant. To capture state effects, we estimated two additional models. For metabolic state, it included the transformed run counter, the metabolic state (hunger–satiety / 100), whether participants consumed caffeine in the last two hours, and whether they had a BE episode. For mood state, it included the transformed run counter, the mood state (happy–sad / 100), and alertness, all as random effects by ID. All priors for parameters concerning the mean were normal distributions with an SD of 1. Priors for variabilities were Cauchy distributions with location 0 and scale 1. For correlations, we used the lkj prior. We fit the models using 8 chains with 4000 iterations, of which 2000 were burn-in. Convergence was assessed using *R̂* ≤ 1.05. To test whether individual differences in variance in learning parameters across runs can be explained by state effects, we correlated the variances estimated in the STAN model with mood and metabolic states, and meta-learning effects extracted from the location scale models (i.e., the random effects of each participant).

#### Single-item analysis of eating behavior

To extract dimensions reflecting inter-individual differences in eating behavior, we used non-negative matrix factorization (NNMF). In contrast to other dimension reduction methods, such as principal component analysis or factor analysis that seek to capture sparse latent representations, NNMF builds an additive, parts-based representation. These parts arise due to non-negativity constraints that eliminate subtractive combinations, thereby facilitating an intuitive interpretation of the identified building blocks of a multidimensional construct such as eating ^75^. It is instructive to consider the data as a matrix within a matrix factorization framework. The questionnaire data is treated as an n (participants) × m (items) matrix, *V*. Each column contains *n* non-negative item scores of one of the *m* items that belong to the six eating questionnaires. NNMF then constructs approximate factorizations of the form *V* ≈ *W*×*H*. The columns of *W* are features (equivalent to factor scores) whereas the column of H are loadings corresponding to the items in *V*. Therefore, the product *W*×*H* is a compressed form of the observed questionnaire data in *V*. In other words, NNMF is a method that models the generation of observable (manifest) variables, *V*, from hidden variables, *H*, where each hidden variable *H* coactivates a subset of observable variables ^75^. In contrast to NNMF, principal component analysis constrains the columns of *W* to be orthonormal, while the rows of *H* are constrained to be orthogonal to each other. Although the resulting eigenvectors have a clear statistical interpretation due to their link to explained variance, they are often not straightforward to interpret ^75^. Notwithstanding, a more mechanistic understanding of the essential parts that contribute to behavioral changes in obesity requires a well-interpretable grounding in manifest eating behavior.

To ensure sufficient stability of the NNMF results (i.e., correlations of identified dimensions across reruns ≥ .92), we used 50,000 replicates in an alternating least squares algorithm with the maximum number of iterations set to 150. All 133 items were rescaled to increase the comparability of loadings across items (0 corresponded to the minimum, 1 to the maximum that we observed in the data).

#### Associations of learning parameters with BMI and binge eating

First, we assessed associations of learning performance (i.e., average points won) with BMI and BE using bootstrapped partial correlations correcting for age and sex. To keep the sample size as large as possible (not all participants in the online game completed the phone screening), we quantified BE as the average number of runs in which a BE episode was reported since the last game in the app (BE EMA). In addition, to isolate the association with either BMI or BE EMA beyond the other, we also corrected for BE EMA in correlation with BMI and the other way around. Second, since learning parameters are often correlated, we used multivariate prediction models to determine a) whether the combination of reward learning parameters explains a significant portion of variance and b) which learning parameters or eating behavior dimensions contribute to BMI and self-reported BE during the EMA phase. To this end, we used an elastic net regression lasso function in MATLAB with the hyperparameter α set to 0.5 a priori ^76,77^. Elastic net is a regularized regression technique combining *L_1_* and *L_2_* penalties used in LASSO and ridge regression, respectively. By selecting a limited number of complementary predictors from a candidate set of correlated features, it generates a sparse model. Elastic net typically outperforms other feature selection methods, such as stepwise regression ^78,79^, and performs well if predictors are highly correlated ^80^. It also performs well in terms of error control in moderately large samples (N∼400) if the number of predictors is low, even if the accuracy continues to increase in larger samples ^79,81^. We used a nested 10-fold cross-validation procedure, where the hyperparameters are always optimized in the inner loops while the outer cross-validation guards against over-fitting and increases generalizability of the results ^82^. We used permutation tests comparing the result against a model with shuffled predictors but still including unshuffled confounders (age, sex, and BMI or BE EMA depending on the outcome) to quantify how much variance learning parameters explain beyond demographic confounders ^76^.

We used the same approach of combining bootstrapped partial correlations and elastic net predictions to determine the association of learning parameters with the transdiagnostic dimensions of eating behavior (derived from the NNMF) that were most predictive of either BMI (i.e., negative emotional eating and cognitive restraint) or self-reported BE EMA (i.e., loss of control and craving). However, in those models we only corrected for age and sex, but not BMI or BE EMA to capture transdiagnostic associations across participant with a high BMI and/or BE.

### fMRI procedure and task

To collect additional lab-based measures of reward, a subsample of 61 participants was invited for additional visits including an fMRI session. During this fMRI session, they performed several tasks including an effort allocation task (EAT) to collect behavioral and neural measures related to reward and cost sensitivity as part of an instrumental motivation task that required no learning of contingencies. The sample included only women with and without BE and consisted of three age and BMI-matched groups: 21 women fulfilling the criteria for BED, 20 women with subsyndromal BE, and 20 women without subjective or objective BE episodes matched for age and BMI to the BE groups. For 59 participants (BED: *n* = 20, *M*_Age_ = 42.8 years ± 12.8, *M*_BMI_ = 33.6 kg/m^2^ ± 6.6; subBED: *n* = 19, *M*_Age_ = 38.3 years ± 12.4, *M*_BMI_ = 28.7 kg/m^2^ ± 7.5; no BE: *n* = 20, *M*_Age_ = 39.1 years ± 14.5, *M*_BMI_ = 32.1 kg/m^2^ ± 6.7), complete data of the EAT is available (2 participants did not complete the task and pressed the alarm ball).

For the fMRI session, participants came in the morning after an overnight fast. After the placement of an intravenous catheter and first blood draws to assess fasting levels of hormones, they received a standardized breakfast (∼350 kcal). The fMRI session started about one hour after the meal and included an anatomical scan, a food bidding task, and the EAT. In the EAT, participants had to collect food and money tokens throughout the task by exerting effort using a grip force device. The task was adapted from Meyniel et al. ^83^ and previous work of our group ^63,84^ now using grip force to measure physical effort. Moreover, instead of two fixed difficulty levels, the task used graded difficulty levels (64 – 95% of individual maximum effort) and an uncertainty condition where the exact amount of effort necessary was not shown. Every trial started with the presentation of the reward at stake for 1 s. The prospective reward could be either food (indicated by cookies) or money (indicated by coins). Moreover, we varied the magnitude of the prospective rewards (1 point vs. 10 points), the difficulty of a trial (ranging from 64 – 95% individual maximum effort), and the uncertainty about the difficulty of a trial. Next, a tube containing a blue ball appeared on the screen with a red line (certain) or red area (uncertain) indicating the level above which the ball had to be moved by squeezing the grip force device to earn reward points. For every second that the ball was kept above the threshold, reward points were accumulated. This led to 64 trials with 24 s effort phases in each trial. At the end of the session, tokens were exchanged for calories in the form of a snack and money. The task was presented using Psychophysics toolbox v3 ^85,86^ in MATLAB v2018a.

### fMRI data acquisition and preprocessing

Functional imaging data was acquired on a Siemens 3T Prisma scanner with a 64-channel head coil. The task lasted approximately 35 minutes and fMRI data during the task were 1,500 T2*-weighted images echo planar images acquired with a multiband sequence (factor 4) with a short TR of 1.4s and TE of 30 ms (flip angle = 65°, field of view = 220 x 220 mm^2^) and voxel size of 2 x 2 x 2 mm^3^. Additionally, an anatomical T1-weighted image was acquired using a MP-RAGE sequence (176 slices, flip angle = 9°, matrix size = 256 x 256, voxel size = 1 x 1 x 1 mm^3^) as well as a field map to correct for field inhomogeneities (TE_short_ = 5.19 ms and TE_long_ = 7.65 ms). Images covered the whole brain and were acquired with 68 slices with an interleaved slice order. Data was then preprocessed using fMRIprep ^87^, including slice time correction, realignment, bias field correction, normalization to MNI space, and segmentation of the anatomical T1 image (fmriprep output, see SI). For confound correction in first-level analysis, we extracted average white matter and CSF signal as well as six motion regressors and framewise displacement of each volume.

#### fMRI data analysis

fMRI data were analyzed using SPM12. The first-level general linear models included regressors for the presentation of the rewards (cue phase), work phases, and feedback phases. Cue and feedback phases were modeled as events (zero duration), and work phases were modeled as blocks with a duration of 24 s. In addition to the task regressors, we included parametric modulators for magnitude (cue and feedback phase) and difficulty (feedback phase). For each TR, we also modeled regressors tracking the exerted force and the change in exerted force. We included the six movement parameters and average white matter and CSF signal as nuisance covariates. Data was high-pass filtered with a cut-off of 128 s.

To measure reward sensitivity independent of learning, we used the change in brain activation in the nucleus accumbens (NAcc) associated with a 10-fold increase in reward magnitude during the cue phase of the EAT (captured in the parametric modulation by reward magnitude). Such measures have been previously used to indicate reward sensitivity ^88–91^ in association with key differences in reward function ^92^. To test for group differences, we extracted the average beta values for each condition (food and money) using the bilateral NAcc region of interest in the Harvard-Oxford brain atlas ^93^ and applied linear mixed-effects model including reward type (as random slope per ID) and age, BMI, and BED status as fixed effects.

#### Statistical threshold and software

For statistical analyses, we used a two-tailed *α* ≤ .05 as the significance threshold. Data for the analyses were processed with MATLAB v2022a and R v4.4.2 ^94^. We implemented NNMF (nnmf function) and elastic net prediction models in MATLAB (lasso function with alpha set a priori to 0.5). Partial correlations were calculated and bootstrapped in MATLAB using *bootci*. Bayesian models for the reinforcement learning data analyses were coded in R ^94^ with the packages *bayestestR* ^95^, *shinystan* ^96^, *tidybayes* ^97^, and *bayesplot* ^98^. Results were plotted in R using *ggplot2* ^99^ and *ggdist* ^97^.

## Results

To characterize eating behavior beyond BMI and self-reported BE (i.e., BE EMA), we extracted transdiagnostic dimensions using NNMF from 133 items. A model with 7 dimensions provided the best trade-off as additional explained variance leveled off after that (see SI). The dimensions capture trait-like eating behaviors ranging from healthy behavior (e.g., eating in response to positive emotional states, or reflective eating) to pathological behavior e.g., loss of control, or craving as dimensions of uncontrolled eating ^100,101^. Accordingly, BMI and BE EMA showed distinct associations with dimensional eating behavior. Whereas BMI was strongly associated with negative emotional eating (*r*_partial_=.40, *p*_boot_<.0005), craving (*r*_partial_=.32, *p*_boot_<.0005), and negatively with reflective eating (*r*_partial_=.-28, *p*_boot_<.0005), BE EMA was strongly associated with craving (*r*_partial_=.51, *p*_boot_<.0005) and loss of control (LoC, *r*_partial_=.40, *p*_boot_<.0005). To determine which dimensions explained differences in BMI or BE EMA best, we used multivariate elastic net predictions and successfully predicted both BMI (*ΔR^2^*=.19; *p_perm_*<.0001) and BE EMA (*ΔR^2^*=.29; *p_perm_*<.0001). These models explained 19% or more of the variance of the outcome (BMI or BE EMA) compared to confounders and the other outcome (BE EMA or BMI, respectively). We then identified the most important features using leave-one-dimension-out predictive models, which pointed for BMI to negative emotional eating (*ΔR^2^*=.049), craving (*ΔR^2^*=.013), reflective eating (*ΔR^2^*=.004), and cognitive restraint (*ΔR^2^*=.014). For BE EMA, the dimensions loss of control (*ΔR^2^*=.036) and craving (*ΔR^2^*=.077) were most predictive. To summarize, BMI and BE EMA were characterized by differential profiles in terms of trait-like transdiagnostic eating behavior that may be associated with distinct learning profiles.

**Figure 2:**
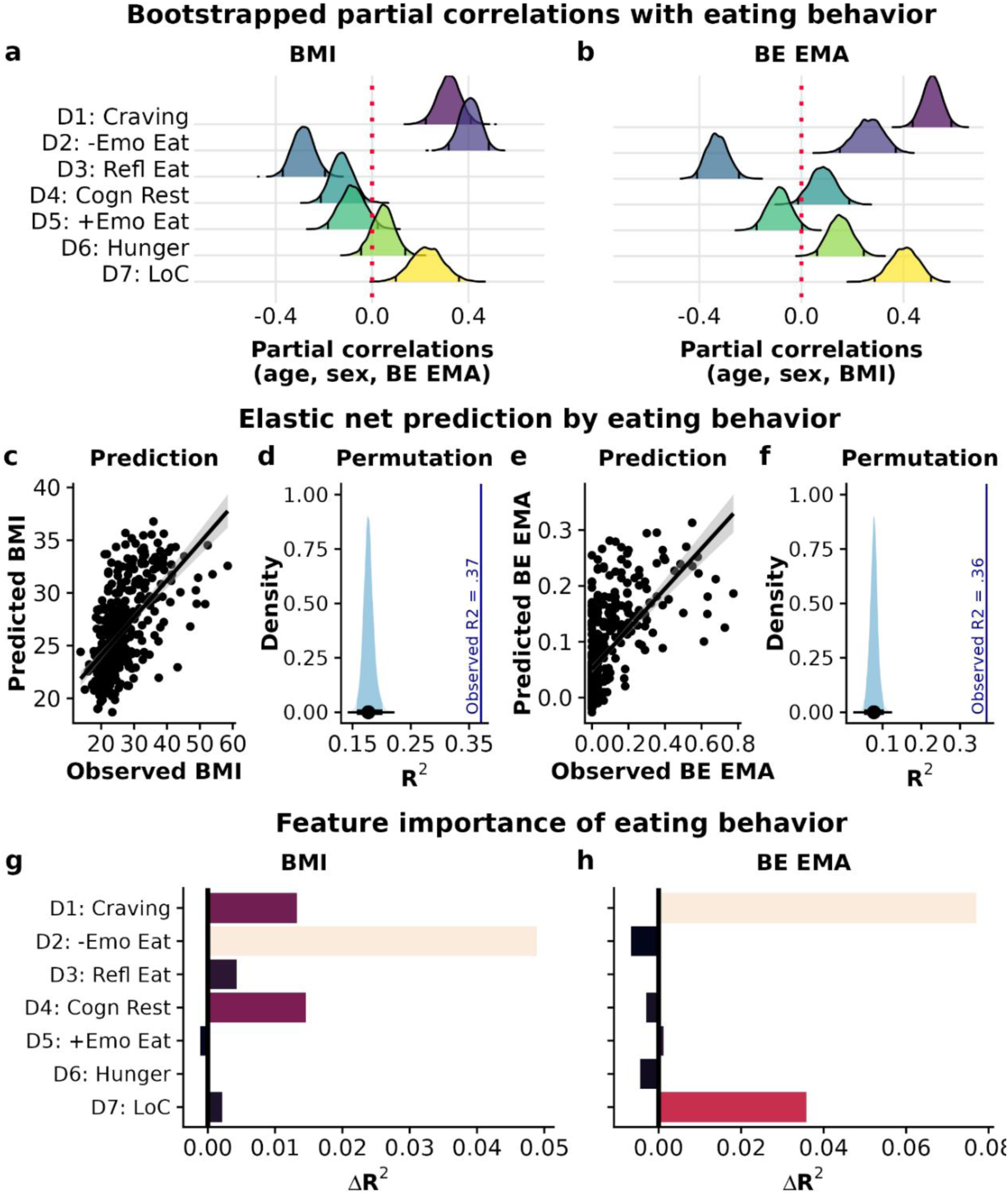
Body mass index (BMI) and self-reported binge eating (BE EMA) are characterized by discernible patterns of trait-like eating behavior. a) Partial correlations corrected for age, sex, and BE EMA show significant positive associations of craving, negative emotional eating, and loss of control as well as negative associations for reflective eating and cognitive restraint with BMI. b: Partial correlations corrected for age, sex, and BMI show significant positive associations of craving, negative emotional eating, eating in response to hunger, and loss of control, as well as negative associations for reflective eating with BE EMA. Distributions (a and b) show bootstrapped (5,000 resamples) partial correlations with 2.5 and 97.5 percentiles. c,e: Models including dimensions of eating behavior predict BMI and BE EMA. Predicted and observed values of BMI and BE EMA were significantly correlated, and the model explained 37% and 36% of the total variance, respectively. d,f: Adding dimensions of eating behavior improves the prediction of BMI and BE EMA compared to the confounding variables. Error bars depict 95% percentiles. g-h: Feature importance of the dimensions of eating behavior for BMI and BE EMA. The most important features contributing to the predictions include negative emotional eating (BMI), craving, and loss of control (BE EMA). The ΔR2 reflects how much predictive accuracy is lost when leaving out the dimension as a feature. -Emo Eat = Negative emotional eating, Refl Eat = Reflective eating, Cogn Rest = Cognitive restraint, +Emo Eat = positive “homeostatic” emotional eating, LoC = loss of control.

### BMI is associated with reward learning parameters

To investigate whether BMI is associated with altered reward learning, we modeled reinforcement learning data from a bandit task with fluctuating win probabilities implemented as a smartphone game. Participants completed the task for between 10 and 31 runs with 150 trials per run (n=10,037 runs included across participants). We then used hierarchical Bayesian models with three levels (M_3L_; run, participant, group) to derive reinforcement learning parameters (mean and variance) for each participant across all of their completed runs ^51^. This model estimating distributions of all parameters for each individual across runs fitted the data better than a model (M_2L_) without the nested data structure (i.e., sampling the parameters of each run directly from one group level prior; ΔWAIC_M2L-M3L_ = 4096.9, ΔLOOIC_M2L-M3L_ = 5837.4) or a model with the nested data structure for β only (ΔLOOIC_M2L.A-M3L_ = 3494.3). This demonstrates that variability across runs differs between participants and that the less complex model leads to stronger shrinkage of individual differences (Fig. 3).

**Figure 3:**
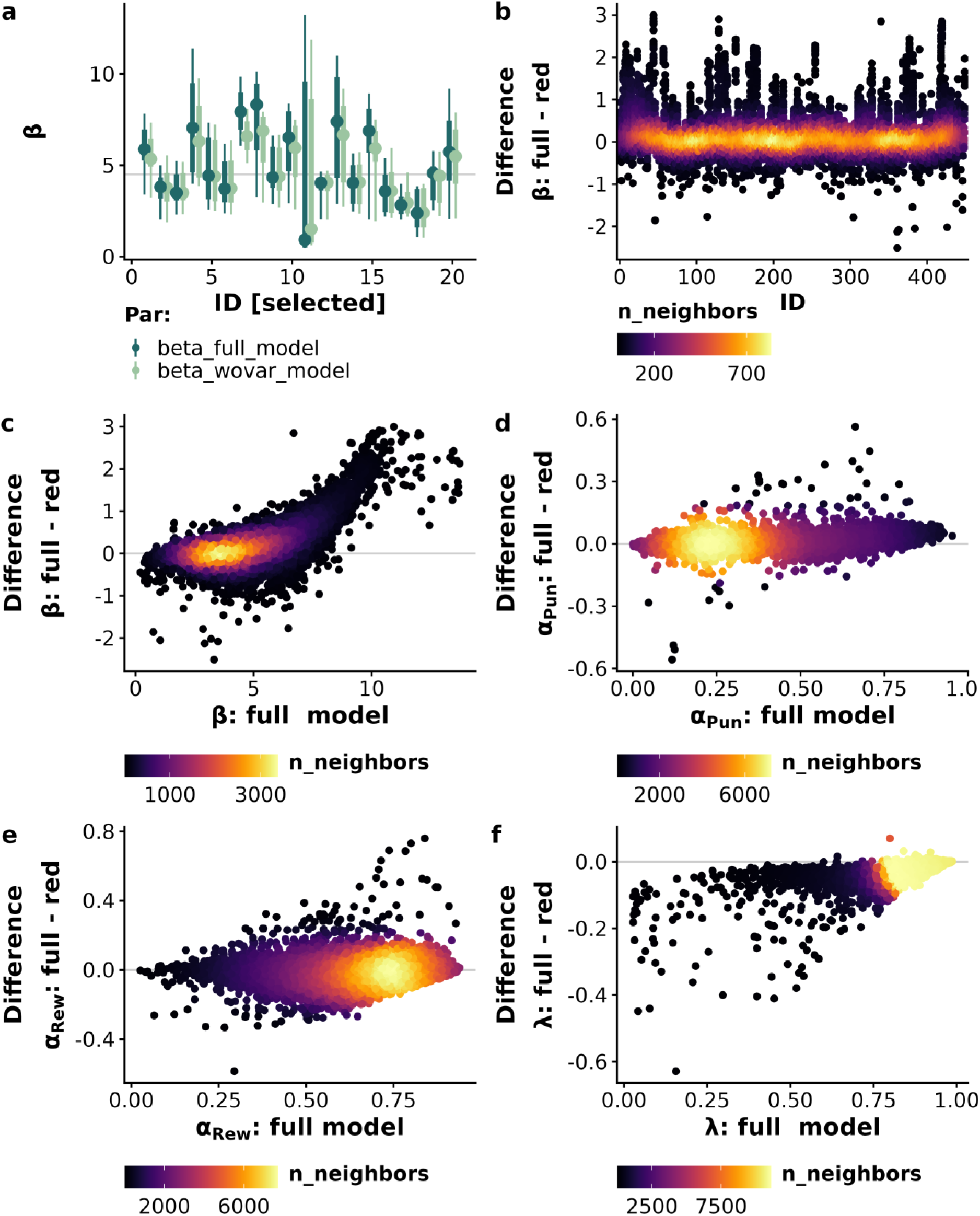
Using the full hierarchical model with individually estimated variability finesses the shrinkage of estimates by incorporating individual differences in behavioral variability. a: Difference in estimated mean and variance of representative IDs for the reward sensitivity β for the full model (dark green) and model without distributions at the individual level (light green). b: Difference between the run estimates for reward sensitivity of the full model – the reduced model sorted by ID. c-e: The full model shows reduced shrinkage for larger (beta) or smaller (lambda) values as individual distributions allow shifts. Shown is the difference (full – reduced model) of all run estimates depending on the estimate in the full model.

Next, we used partial correlations to test the link between learning performance and BMI and found no association of average reward with BMI (*r*_partial_=-.04, *p*_boot_=.46). Since different parameter combinations may lead to similar performance, we used cross-validated elastic net regression based on learning parameters to predict BMI beyond confounding variables including BE EMA (*ΔR^2^*=.030; *p*_perm_=.007). In this model, higher average learning rates for rewards and punishments, as well as higher variabilities contributed to the prediction of BMI (Fig. 3e). Next, we evaluated whether transdiagnostic eating behavior associated with BMI was similarly explained by altered learning behavior, but negative emotional eating and craving were not successfully predicted by learning parameters (Table S1; *p*s > .27), although craving was associated with fewer points won (*r*_partial_=-.12, *p*_boot_=.036, Fig. S3).

**Figure 3:**
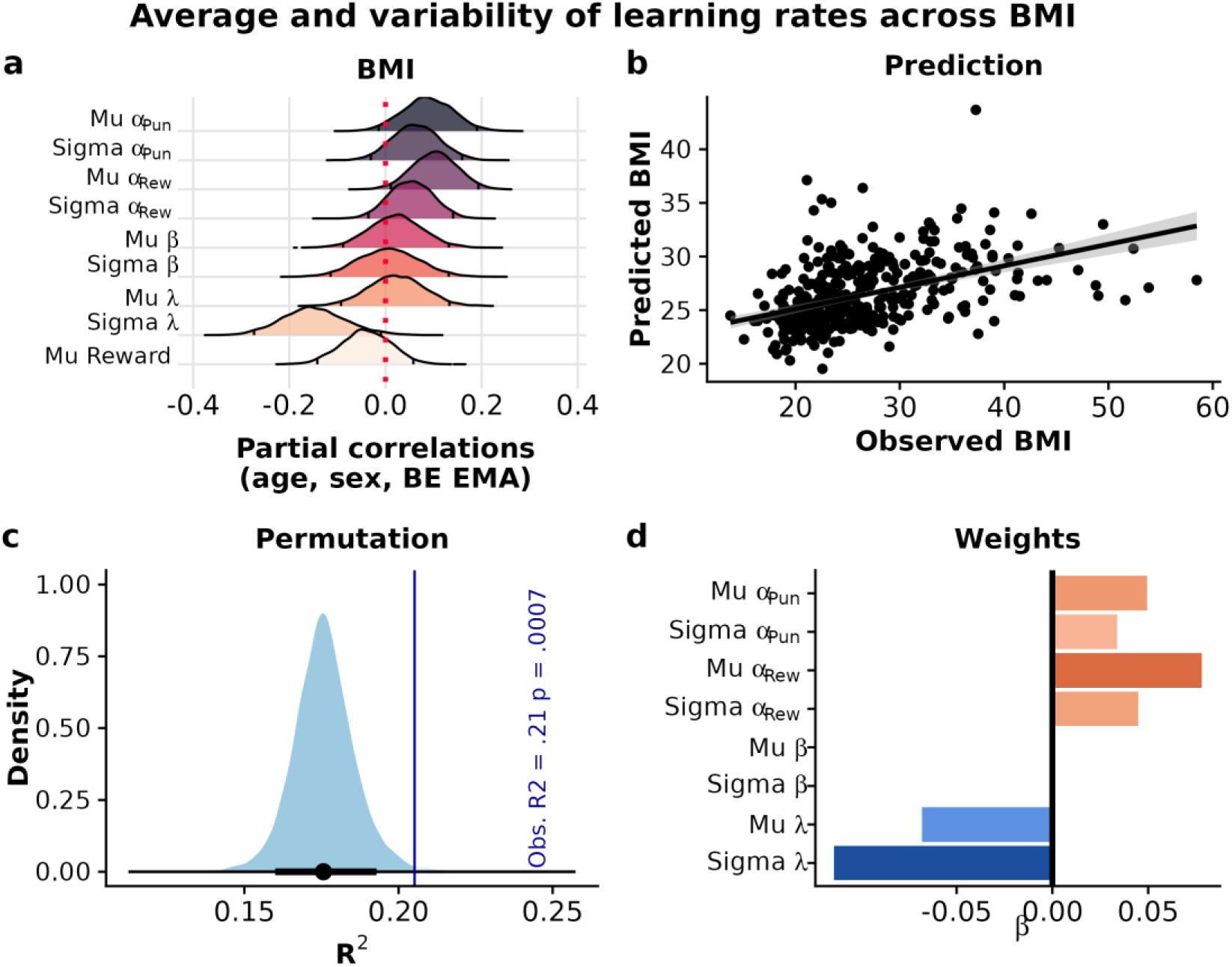
Average learning rates for rewards and punishments and their variability across runs are associated with body mass index (BMI). a: Partial correlations corrected for age, sex, and self-reported binge eating (BE EMA) show positive associations of BMI with higher learning rates for rewards and lower variability in the use of reward magnitude to guide decisions (i.e., λ). Variability estimates are residualized for differences in the average and reflect only incremental variance. Distributions show bootstrapped (5,000 resamples) the partial correlations with 2.5 and 97.5 percentiles. b: A cross-validated elastic net model predicted BMI using learning parameters. Predicted and observed values of BMI were correlated and the model explained 21% of the total variance. c: Adding reinforcement learning parameters improves the prediction of BMI compared to the confounding variables of age, sex, and BE EMA. Error bars depict 95% percentiles. d: Feature weights contributing to the prediction of BMI. Higher average learning rates and their variability together with lower average λ and its variability contribute to the prediction.

### Binge eating episodes and loss of control are associated with reward sensitivity

To disentangle learning differences associated with self-reported BE versus BMI, we applied the same analyses with BE EMA as the outcome and BMI as an additional covariate. In contrast to BMI, BE EMA was associated with fewer earned points per run (*r*_partial_=-.14, *p*_boot_=.022, Fig. 4a). To determine whether specific patterns of learning behavior underlie the reduced performance, we again used cross-validated elastic net regression. We found that BE EMA was predicted by a distinct set of features compared to BMI including a lower average reward sensitivity and higher variabilities for reward sensitivity, learning rates for rewards, and the use of reward magnitude to guide decisions (*ΔR^2^*=.023; *p_perm_*=.024; Fig. 4d).

**Figure 4:**
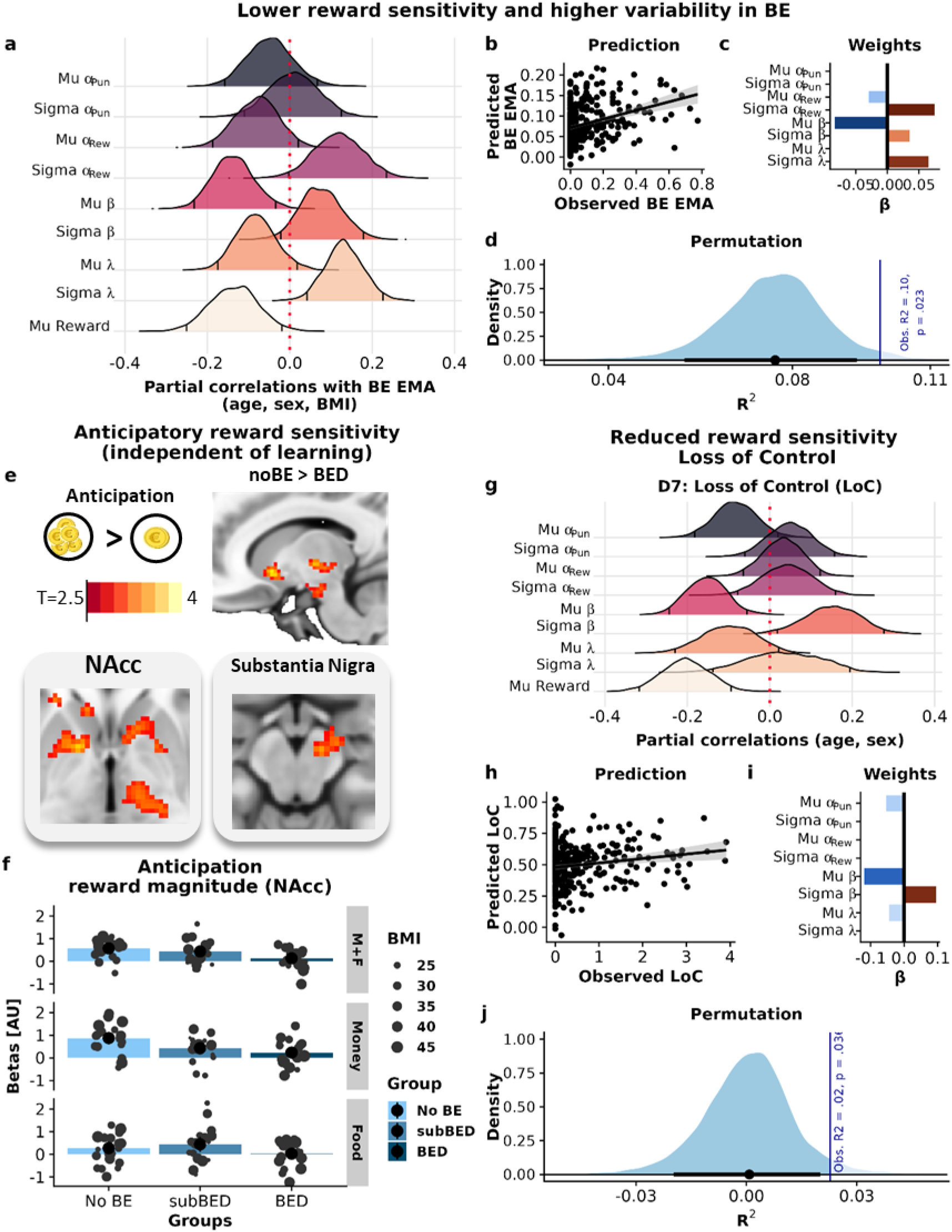
Binge eating (BE) is associated with reduced but more variable reward sensitivity. a: Partial correlations corrected for age, sex, and body mass index (BMI) show associations of BE EMA with fewer earned points, lower average reward sensitivity, and higher variability in use of reward magnitude to guide choices (i.e., λ). Variability estimates are residualized for differences in the average. Distributions show bootstrapped (5,000 resamples) partial correlations with 2.5 and 97.5 percentiles. b: A model including learning parameters predicted BE EMA. Predicted and observed values of BE EMA were correlated and the model explained 9% of the variance. c: Feature weights contributing to the prediction of BE EMA. Lower reward sensitivity and learning rates for rewards, together with higher variability of learning rates for rewards, reward sensitivity, and λ contribute to the prediction. d: Adding reinforcement learning parameters improves the prediction of BE EMA compared to the confounding variables age, sex, and BMI. Error bars depict 95% percentiles. e-f: Anticipatory brain responses in the nucleus accumbens (NAcc) for high vs. low reward magnitude are reduced in patients with BE disorder across all trials (*t*=--2.86, *p*=.006) and independent of reward type (Food x BED: *t*=1.44, *p*=.15). g: The dimension loss of control (LoC) is associated with fewer earned points, together with lower and more variable reward sensitivity using partial correlations corrected for age and sex. Distributions show bootstrapped (5,000 resamples) partial correlations with 2.5 and 97.5 percentiles. h: A model including learning parameters predicted LoC. Predicted and observed values of LoC were correlated, and the model explained 2.2% of the variance. i: Feature weights contributing to the prediction of LoC. Lower and more variable reward sensitivities contribute to the prediction. I: Adding reinforcement learning parameters improves the prediction of LoC eating compared to confounds.

In reward learning tasks, it is challenging to clearly dissociate learning rates and reward sensitivity (i.e., scaling of the subjective value) because the parameters are correlated ^102^. To evaluate if reduced reward sensitivity in BE generalizes to other tasks that require no learning, we analyzed fMRI data from a subset of 59 participants with vs. without BED (age and BMI-matched). In the EAT, participants worked to earn food or monetary rewards using a grip force device. To capture reward sensitivity, we extracted the NAcc response during the anticipation phase of a trial and compared high vs. low reward magnitude trials (10 vs. 1 points per second). Likewise, we estimated behavioral reward sensitivity as the individual adjustment in effort for high vs. low reward magnitude trials (i.e., individual slopes of a linear mixed-effects model). In line with the interpretation that the average β reflects reward sensitivity, we observed a correlation with the increase of effort for high-magnitude rewards (*r*(56)=.31, *p*=.018, Fig. S4). Moreover, participants with BED showed a reduced NAcc response to higher reward magnitude (*b*_BED_=-0.43, *t*(53)=-2.86, *p*=.006, Fig. 4f), whereas BMI was not associated with NAcc responses (*t*(53)=0.42, *p*=.67, model controls for age). This reduced NAcc reward sensitivity was not specific for food rewards in patients with BED (BED x Food: *t*(56)=1.44, *p*=.15), suggesting a generally reduced reward sensitivity independent of learning. Notably, NAcc anticipatory NAcc responses were associated with the effort allocated in the subsequent trial in food trials (*t*(53)=2.02, *p*=.049).

To determine if reduced and more variable reward sensitivity is captured by transdiagnostic dimensions of eating behavior characterizing BE (i.e., loss of control and craving), we again used cross-validated elastic net models but only accounted for sex and age. Analogous to BE EMA, loss of control was predicted by reduced and more variable reward sensitivity as well as lower punishment learning rates and *λ* (*ΔR^2^*=.022; *p_perm_*=.036, Fig. 4j). Moreover, a higher loss of control was also associated with a significantly lower number of points per run (*r*_partial_=-.21, *p*_boot_<.001). Notably, accounting for loss of control in the partial correlations between learning parameters and BE EMA attenuated the associations of BE EMA with reward sensitivity (average: *r*_partial_=-.08, *p*_boot_=.13 and variability: *r*_partial_=.01, *p*_boot_=.81) and earned points (*r*_partial_=-.06, *p*_boot_=.33; Fig. S5), indicating that these differences in BE are partially mediated by loss of control. In contrast to loss of control, craving was not associated with differences in learning behavior.

### Variability in reinforcement learning is partly explained by mood and metabolic states

Since BMI and BE EMA were associated with elevated variability in learning rates or reward sensitivity, this raises the question of whether this is due to differences in mood or metabolic states. To answer this question, we capitalized on the concurrent EMA questions and estimated the individual-level effects of mood state and metabolic state on learning parameters using mixed-effects models in brms (while accounting for increased task performance over runs). Higher individual variability in all parameters was associated with larger individual improvements over runs (*p*s<.0001, Fig. 5c,f,i). In addition, variability in learning rates for punishments and reward sensitivities were higher in participants with larger mood-related increases (i.e., *r*_β_ = .40, *r*_αPun_ = .31, *p*s<.005). In contrast, participants with higher variability in learning rates for wins, showed a reduced mood-related increase (*r*_αRew_ = −.27, p<.001). In addition to mood state, adaptations to the current metabolic state (‘metabolic scaling’) were associated with variabilities in reward sensitivity (*r*=.42, *p*<.001) and learning rates for punishments (*r*=-.41, *p*<.001), but not learning rates for wins (*r*=.07, *p*=.19). However, in participants with higher BMI variability of learning rates for wins were also associated with metabolic scaling (BMI x SD: *b*=0.002, *p*<.001, Fig S6). Notably, metabolic scaling of learning rates was reduced for participants with a higher BMI (*r*_αPun_ = −.13, *p*_αPun_ =

**Figure 5:**
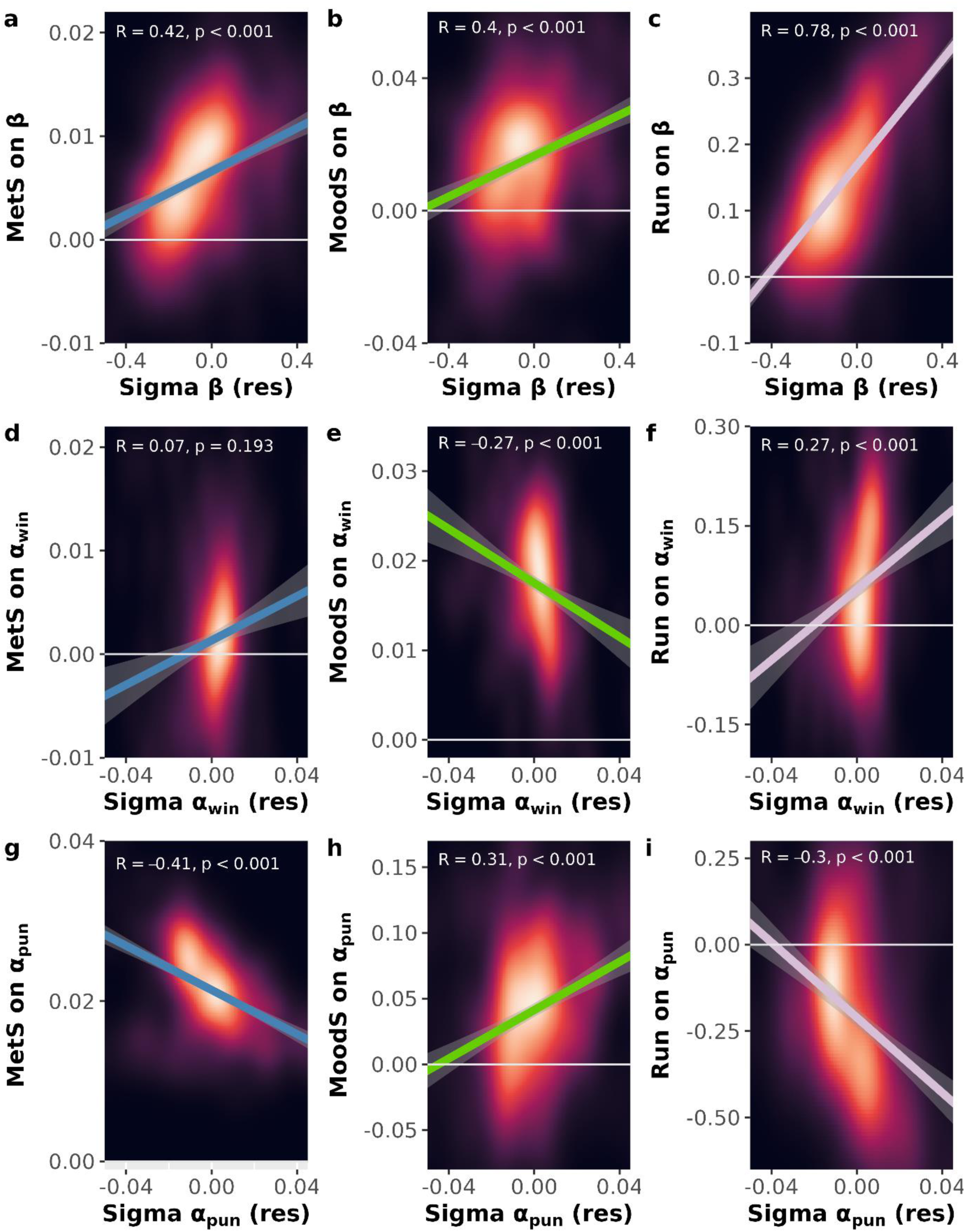
Variability in reinforcement learning (RL) parameter estimates is partly explained by improvements over runs and state effects of mood and metabolism. Correlations between variabilities of all parameters (residualized for the mean, for lambda see Fig. S7) and individual effect estimates (individual slopes extracted from the multilevel models) for changes over runs (Run), mood state (MoodS), and metabolic state (MetS) modulating learning parameters. The color gradient of the background shows the density of observations ranging from black, indicating low density, to white, which indicates a high density of observations.

.010, *r*_αWin_ = −.18, *p*_αWin_ <.001). In other words, participants with overweight and obesity did not learn more quickly from rewards or punishments when they were hungry. Comparably, metabolic scaling of reward sensitivity was elevated in participants reporting higher loss of control (*r*_beta_ = .14, *p*_beta_ = .011). Together, these results show that differences in the adjustment of reinforcement learning by internal states partly explain differences in behavioral variability and contribute to eating-related pathologies.

### Hedonic eating is associated with changes in learning

Beyond transdiagnostic eating dimensions associated with pathological behavior, we explored if dimensions of healthy eating behavior (i.e., positive emotional eating or “homeostatic” eating in response to hunger), also showed distinct profiles of learning behavior. Trait-like positive emotional eating was associated with more earned points (*r*_partial_=.11, *p*_boot_=.032). Consequently, cross-validated elastic net models successfully predicted positive emotional eating (*ΔR^2^*=.021; *p_perm_*=.033, Fig. S8) and the identified pattern of learning behavior (lower α_pun_, higher β and λ) resembles the behavior of optimal agents on the task, explaining why it is associated with more earned points.

## Discussion

Although obesity and eating disorders are characterized by changes in dopamine signaling, the evidence for altered reward learning in obesity is mixed and it is unclear whether obesity and comorbid eating disorders (e.g., BED) show distinct learning profiles. Here, we use a dense sampling approach of choices in a gamified learning task to show that obesity and BE are associated with differential changes in learning behavior. Obesity was characterized by increased learning rates that did not translate to reduced performance in our task. In contrast, BE and transdiagnostic loss of control were characterized by reduced wins reflected in lower and more variable reward sensitivity in a reinforcement learning task. Crucially, in patients with BED, this was mirrored by reduced NAcc activation in anticipation of larger rewards, quantifying reward sensitivity independent of learning or decision noise. This alignings with the reward deficit hypothesis of BE ^103^ and is in contrast with the common hypothesis that reward sensitivity is increased in BED ^46^ based on non-instrumental tasks suggesting that behavioral outcomes are essential for the understanding of BE. Remarkably, our densely sampled choice data over weeks further revealed that across runs learning behavior was more variable in both obesity (i.e. higher variability in learning rates) and BE (i.e., higher variability in reward sensitivity) and that this variability was associated with internal affective and metabolic states. This highlights the importance of assessing state-dependent fluctuations in behavior to unravel mechanisms that characterize distinct types of pathological eating behavior. To conclude, our study shows that an in-depth phenotyping of the behavioral signatures across states is necessary for mechanistic insights into obesity and eating-related psychopathology.

In contrast to one common theory that BED is characterized by higher reward sensitivity ^46^, we find that reward sensitivity was reduced supporting the opposing view that BED is characterized by a reward deficit ^103^. Rewards sensitivity was reduced, both in the learning task and an instrumental effort task comparing NAcc activity in anticipation of rewards, and there was no difference between food and monetary rewards. This adds evidence to a reward deficit account of BE compared to a reward surfeit hypothesis postulated for obesity and weight gain ^103^. Many previous studies rely on the passive viewing of (food) reward pictures or questionnaires to quantify reward sensitivity, whereas we analyzed data from instrumental tasks with and without a learning requirement. Our findings are in line with previous work that showed lower reward sensitivity (or an increase in stochasticity of choices) in patients with BED ^39,47–49^. Whereas it is often hard to distinguish lower reward sensitivity from noisier decision-making in a reinforcement learning paradigm ^104^, our study provides evidence against this. Crucially, reward sensitivity was also reduced in anticipation of rewards during the EAT, which does not include learning or decision noise components. Moreover, as hypothesized, reward sensitivity and the learning rate for wins were more variable in participants reporting more BE episodes. Although a higher variability in reward sensitivity across runs may also be explained by more noise, the large number of runs allowed us to estimate state effects in addition to noise. There, an increase in variability was related to stronger adaptations depending on mood and metabolic states ^105–107^. Notably the metabolic scaling of the learning rates lower with a higher BMI, in line with a reduced effect of hunger on motivation in participants with obesity^108^. In contrast, loss of control was associated with a larger metabolic scaling of the reward sensitivity, supporting the idea that larger excursions from energy homeostasis may contribute to the risk for BE ^109–111^ comparable to periods of more negative mood ^53,112–114^. Moreover, metabolic signals from the gut potentially shape learning^115^ and goal-directed behavior via altered dopamine transmission ^40,63,116–119^. The transdiagnostic dimension ‘loss of control’ accounted for lower and more variable reward sensitivity and lower average rewards suggesting that extreme states of reward sensitivity and phases of loss of control over eating are linked. Loss of control was not related to the variability in learning rates for wins. Taken together, lower reward sensitivity across tasks and larger behavioral variability across states characterize increased BE and loss of control over eating.

As hypothesized, obesity was associated with changes in learning behavior, although it did not lead to detrimental performance. In contrast to valence-specific predictions, participants with higher BMI showed increased learning rates for both wins and punishments ^27–31^. Remarkably, obesity was not associated with reward sensitivity when accounting for BE, suggesting that an elevated BMI is correlated with faster reinforcement learning independent of valence and subjective value. In highly volatile environments, higher learning rates are beneficial, but in comparatively stable environments, including the current task, lower punishment learning rates are optimal according to simulations ^51^. In line with larger variability in patients with obesity, wanting ratings ^120^ and reward responses in the NAcc have been shown to be more variable in obesity across tasks ^34^. Conceptually, elevated learning rates may also reflect a shorter span of reward history that could lead to more exploratory or impulsive behavior ^56,121–123^ and this could be explained by altered dopamine transmission in obesity ^21–26^. Previous reports of changes in reward sensitivity e.g., ^32^ may be partly explained by confounding with BE that we controlled for in our analysis. Likewise, reward sensitivity may have been hard to distinguish from learning itself in learning task. Crucially, our measure of reward sensitivity but not the learning rates are also associated with behavioral reward sensitivity in a task without a learning component. To summarize, our extensive data of 370 participants with more than 1,500 choices per person shows that obesity is associated with higher learning rates, but not changes in reward sensitivity.

Although our study has several strengths (e.g., a large sample with dense sampling of choices), there are limitations that should be considered. First, the completion of the reinforcement learning game and the mood and metabolic self-reports were timed by the participants. Future studies may use specific triggers related to mood or metabolic states to test whether BE or loss of control occurs in extreme states of reward sensitivity. Second, we only assessed self-reported metabolic state, and concurrent “objective” measures derived from continuous glucose monitoring would be beneficial. Nevertheless, our work^124^ suggests that there is a good agreement between metabolic state ratings and glucose levels in metabolically healthy participants. Likewise, controlled manipulations of metabolic states during functional neuroimaging and learning tasks would help resolve the how metabolic state affects brain responses to rewards causally. Third, while we successfully implemented 3-level hierarchical Bayesian models to estimate individual means and variances in parameters across sessions, these models do not include fluctuations in mood or metabolic states, or run effect. We relied on post hoc mixed-effects models in brms because full models did not converge (for details see SI). Fourth, although all participants from the online study completed an in-depth phenotyping with various questionnaires capturing eating behavior, including BE episodes during the EMA phase, we only completed a full diagnostic interview for the subsample doing additional lab-based sessions. Future work may attempt to better differentiate subjective and objective BE episodes. Fifth, our sample included more female than male participants (∼4:1) since BE is much more prevalent in women ^125^. Attempting to include more men in future studies would allow to investigate sex-specific effects.

Obesity and BED are often comorbid and are both characterized by altered dopamine transmission and reward responses, particularly in response to food. In line with the crucial role of the reward system in learning and decision-making, we showed that obesity and BE are associated with discernable changes in learning behavior. Whereas obesity was related to increased learning rates for rewards and punishments, BE was associated with decreased reward sensitivity. Remarkably, obesity and BE showed higher fluctuations over repeated runs in reward sensitivity and learning rates, respectively. These fluctuations were not only driven by noise since individual variability was associated with ratings of mood and metabolic states, highlighting that internal states likely contribute to altered decision-making in both obesity and BE— albeit with distinct neurocomputational signatures of value and learning. Taken together, our results show that dense sampling of choices across diverse subjective states provides novel insights into the pathomechanism of disordered eating by demonstrating that alleged trait-like behavioral differences are strongly contextualized by state-dependent influences.

## Data Availability

The run level data without participant characteristics is available online at https://osf.io/92s4d/files/osfstorage

## Acknowledgment

We thank Dana Wentz, Mechteld van den Hoek Ostende, Jacob Schwab, and Lillith Irtel von Brenndorff for their help with data acquisition. The study was supported by the Else Kröner-Fresenius Stiftung, grant 2017_A67, and the German Research Foundation (DFG), grants KR 4555/7-1, KR 4555/9-1, & KR 4555/10-1. VT & NBK received salary support from the University of Tübingen, fortune grant #2453-0-0. VT is supported by a Government of Ireland Postdoctoral Fellowship (GOIPD/2023/1238).

## Author contributions

NBK was responsible for the study concept and design. JZ, MPN, VT, & FM collected data under supervision by SV and NBK. NBK conceived the method and AK & JZ processed the behavioral data and VT the imaging data. AK, JT, and FK implemented the hierarchical computational model in consultation with PD. AK, JZ & NBK conducted analyses of questionnaires and associations with reward learning. AK & NBK performed analyses of the imaging data. AK & NBK wrote the manuscript. All authors contributed to the interpretation of findings, provided critical revision of the manuscript for important intellectual content, and approved the final version for publication.

