## Supporting Information for "Dense sampling of choices links high learning rates to obesity and low reward sensitivity to binge eating"

Venusberg Campus 1, 53127 Bonn, Germany

### Food questionnaires

To measure dimensional aspects of pathological eating, we used the Three-Factor Eating Questionnaire, TFEQ (Stunkard and Messick, 1985). The TFEQ consists of three subscales: a) cognitive restraint, b) disinhibition, and c) susceptibility to hunger. It contains 51 items (21 for cognitive restraint, 16 for disinhibition, and 14 for hunger). We calculated scores for each subscale by taking the sum of the items. Out of the three subscales, disinhibition and hunger are thought to be associated with uncontrolled eating (Vainik et al., 2019). In contrast, cognitive restraint has been shown to reflect a separable construct that interacts with uncontrolled eating, thereby relating to BMI (Vainik et al., 2019).

To measure the psychological impact of the presence of palatable food in the environment, we used the Power of Food Scale, PFS (Cappelleri et al., 2009). The PFS consists of 15 items, each rated on a 5-point Likert scale (1: I don't agree to 5: I strongly agree). The PFS encompasses three subscales defined by the proximity of food: a) available but not directly present in the environment, b) present but not tasted, and c) tasted but not consumed. We calculated scores for each subscale by taking the average over its items. All subscales of the PFS are associated with uncontrolled eating (Vainik et al., 2015; Vainik et al., 2019).

To measure addiction-like dependence on food (i.e., a strong indication of uncontrolled eating (Vainik et al., 2019)), we used the Yale Food Addiction Scale, YFAS (Gearhardt et al., 2009). The YFAS consists of 25 items. 16 items were rated on a 5-point Likert scale (0: never to 4: 4 or more times a week or daily) and 9 items were answered as dichotomous questions (0: no, 1: yes). We calculated the score by summing over the (recoded) values associated with each answer (Meule et al., 2017).

To measure eating behavior in relation to specific emotions, we used the Salzburg Emotional Eating Scale, SEES (Meule et al., 2018, doi: 10.3389/fpsyg.2018.00088). The SEES consists of four subscales representing one positive and three negative emotions: a) happiness, b) sadness, c) anger, and d) anxiety. Each subscale contains five items, each rated on a 5-point Likert scale (1: I eat much less than usual to 5: I eat much more than usual). We calculated the score for each subscale by taking the average over its items (Meule et al., 2018, doi: 10.3389/fpsyg.2018.00088).

To measure eating behavior in response to stress, we used the Salzburg Stress Eating Scale, SSES (Meule et al., 2018, doi: 10.1016/j.appet.2017.10.003). The SSES consists of ten items, each rated on a 5-point Likert scale (1: I eat much less than usual to 5: I eat much more than usual). We calculated the score by taking the average of all items (Meule et al., 2018, doi: 10.1016/j.appet.2017.10.003).

To measure food cravings, we used the short version of the Food Cravings Questionnaire – Trait, FCQ-T-r (Meule et al., 2014, doi: 10.3389/fpsyg.2014.00190). The FCQ-T-r consists of 15 items, each rated on a 6-point Likert scale (1: never to 6: always). We calculated the sum score over all items (Meule et al., 2014, doi: 10.3389/fpsyg.2014.00190).

### fMRIPrep Preprocsessing Boilerplate

Results included in this manuscript come from preprocessing performed using fMRIPrep 20.1.1 (Esteban, Markiewicz, et al. (2018); Esteban, Blair, et al. (2018);

RRID:SCR_016216), which is based on Nipype 1.5.0 (Gorgolewski et al. (2011); Gorgolewski et al. (2018); RRID:SCR_002502).

#### *Anatomical data preprocessing*

A total of 1 T1-weighted (T1w) images were found within the input BIDS dataset. The T1-weighted (T1w) image was corrected for intensity non-uniformity (INU) with N4BiasFieldCorrection (Tustison et al. 2010), distributed with ANTs 2.2.0 (Avants et al. 2008, RRID:SCR_004757), and used as T1w-reference throughout the workflow. The T1w-reference was then skull-stripped with a Nipype implementation of the antsBrainExtraction.sh workflow (from ANTs), using OASIS30ANTs as target template. Brain tissue segmentation of cerebrospinal fluid (CSF), white-matter (WM) and gray-matter (GM) was performed on the brain-extracted T1w using fast (FSL 5.0.9, RRID:SCR_002823, Zhang, Brady, and Smith 2001). Brain surfaces were reconstructed using recon-all (FreeSurfer 6.0.1, RRID:SCR_001847, Dale, Fischl, and Sereno 1999), and the brain mask estimated previously was refined with a custom variation of the method to reconcile ANTs-derived and FreeSurfer-derived segmentations of the cortical gray-matter of Mindboggle (RRID:SCR_002438, Klein et al. 2017). Volume-based spatial normalization to one standard space (MNI152NLin2009cAsym) was performed through nonlinear registration with antsRegistration (ANTs 2.2.0), using brain-extracted versions of both T1w reference and the T1w template. The following template was selected for spatial normalization: ICBM 152 Nonlinear Asymmetrical template version 2009c [Fonov et al. (2009), RRID:SCR_008796; TemplateFlow ID: MNI152NLin2009cAsym],

#### *Functional data preprocessing*

For each of the 3 BOLD runs found per subject (across all tasks and sessions), the following preprocessing was performed. First, a reference volume and its skull-stripped version were generated using a custom methodology of fMRIPrep. Head-motion parameters with respect to the BOLD reference (transformation matrices, and six corresponding rotation and translation parameters) are estimated before any spatiotemporal filtering using mcflirt (FSL 5.0.9, Jenkinson et al. 2002). BOLD runs were slice-time corrected using 3dTshift from AFNI 20160207 (Cox and Hyde 1997, RRID:SCR_005927). A B0-nonuniformity map (or fieldmap) was estimated based on a phase-difference map calculated with a dual-echo GRE (gradient-recall echo) sequence, processed with a custom workflow of SDCFlows inspired by the epidewarp.fsl script and further improvements in HCP Pipelines (Glasser et al. 2013). The fieldmap was then co-registered to the target EPI (echo-planar imaging) reference run and converted to a displacements field map (amenable to registration tools such as ANTs) with FSL's fugue and other SDCflows tools. Based on the estimated susceptibility distortion, a corrected EPI (echo-planar imaging) reference was calculated for a more accurate co-registration with the anatomical reference. The BOLD reference was then co-registered to the T1w reference using bbregister (FreeSurfer) which implements boundary-based registration (Greve and Fischl 2009). Co-registration was configured with six degrees of freedom. The BOLD time-series (including slice-timing correction when applied) were resampled onto their original, native space by applying a single, composite transform to correct for head-motion and susceptibility distortions. These resampled BOLD time-series will be referred to as preprocessed BOLD in original space, or just preprocessed BOLD. The BOLD time-series were resampled into standard space, generating a preprocessed BOLD run in MNI152NLin2009cAsym space. First, a reference volume and its skull-stripped version were generated using a custom methodology of fMRIPrep. Several confounding time-series were calculated based on the preprocessed BOLD: framewise displacement (FD), DVARS and three region-wise global signals. FD was computed using two formulations following Power (absolute sum of relative motions, Power et al. (2014)) and Jenkinson (relative root mean square displacement between affines, Jenkinson et al. (2002)). FD and DVARS are calculated for each functional run, both using their implementations in Nipype (following the definitions by Power et al. 2014). The three global signals are extracted within the CSF, the WM, and the whole-brain masks. Additionally, a set of physiological regressors were extracted to allow for component-based noise correction (CompCor, Behzadi et al. 2007). Principal components are estimated after high-pass filtering the preprocessed BOLD time-series (using a discrete cosine filter with 128s cut-off) for the two CompCor variants: temporal (tCompCor) and anatomical (aCompCor). tCompCor components are then calculated from the top 5% variable voxels within a mask covering the subcortical regions. This subcortical mask is obtained by heavily eroding the brain mask, which ensures it does not include cortical GM regions. For aCompCor, components are calculated within the intersection of the aforementioned mask and the union of CSF and WM masks calculated in T1w space, after their projection to the native space of each functional run (using the inverse BOLD-to-T1w transformation). Components are also calculated separately within the WM and CSF masks. For each CompCor decomposition, the k components with the largest singular values are retained, such that the retained components' time series are sufficient to explain 50 percent of variance across the nuisance mask (CSF, WM, combined, or temporal). The remaining components are dropped from consideration. The head-motion estimates calculated in the correction step were also placed within the corresponding confounds file. The confound time series derived from head motion estimates and global signals were expanded with the inclusion of temporal derivatives and quadratic terms for each (Satterthwaite et al. 2013). Frames that exceeded a threshold of 0.5 mm FD or 1.5 standardised DVARS were annotated as motion outliers. All resamplings can be performed with a single interpolation step by composing all the pertinent transformations (i.e. head-motion transform matrices, susceptibility distortion correction when available, and co-registrations to anatomical and output spaces). Gridded (volumetric) resamplings were performed using antsApplyTransforms (ANTs), configured with Lanczos interpolation to minimize the smoothing effects of other kernels (Lanczos 1964). Non-gridded (surface) resamplings were performed using mri_vol2surf (FreeSurfer).

Many internal operations of fMRIPrep use Nilearn 0.6.2 (Abraham et al. 2014, RRID:SCR_001362), mostly within the functional processing workflow. For more details of the pipeline, see the section corresponding to workflows in fMRIPrep's documentation.

#### *Copyright Waiver*

The above boilerplate text was automatically generated by fMRIPrep with the express intention that users should copy and paste this text into their manuscripts unchanged. It is released under the CC0 license.

#### *Prior distributions STAN model*

$$Participant\ estimate: a_{11} \sim \mathrm{Gamma}(2,2)$$

$$Participant\ estimate: a_{12} \sim \mathrm{Gamma}(5,2)$$

$$Participant\ estimate: a_{21} \sim \mathrm{Gamma}(5,2)$$

$$Participant\ estimate: a_{22} \sim \text{Gamma}(4,2)$$

$$Participant\ estimate: b_1 \sim \text{Gamma}(5,1)$$

$$Participant\ estimate: \mu_\beta \sim \text{Normal}(4.5, 2)[0, Inf]$$

$$Participant\ estimate: b_2 = (1 + b1)/\mu_\beta \qquad 1)$$

$$Participant\ estimate:\ l_1 \sim \text{Gamma}(5, 2)$$

$$Participant\ estimate: l_2 \sim \text{Gamma}(1, 3)$$

$$Run\ estimate: \alpha_{loss} \sim \text{Beta}(1 + a_{11}, 1 + a_{12})$$

$$Run\ \ estimate: \alpha_{win} \sim \text{Beta}(1 + a_{21}, 1 + a_{22})$$

$$Run\ \ estimate: \beta \sim \text{Gamma}(1 + b_1, b_2)$$

$$Run\ \ estimate: \lambda \sim \text{Beta}(1 + l_1, 1 + l_1)$$

Here, '~' indicates that parameters are sampled from the stated distribution, and the gamma-distribution is parameterized with a shape and rate parameter (inverse scale), following Stan's modeling language.

#### *Exploration of Hierarchical Bayesian models including state effects*

To estimate the influence of current states (e.g., metabolic state) as well as improvements over runs on reinforcement learning parameters, we initially aimed to extend the hierarchical Bayesian model by run-wise modulations of the parameters. In those models, the effect of a potential modulator (Run number, mood state or metabolic state) was modeled separately for each participant, but the assumed to be constant across time, while the state itself varied between the runs. Accordingly, we added a weight parameter to the model parameters of interest, either the learning rates ($\alpha_{lose}$ and $\alpha_{win}$) or the reward sensitivity $\beta$.

For a given run *i* of the participant *s*, the learning rates could change as follows:

$$\alpha'^{\,i}_{\text{loss}} = \alpha^i_{\text{loss}} + \eta^s_1 * \text{state}^i$$
$$\alpha'^{\,i}_{\text{win}} = \alpha^i_{win} + \eta^s_2 * \text{state}^i \qquad 2)$$

with $\alpha'^{\,i}_{\text{loss}}$ and $\alpha'^{\,i}_{\text{win}}$ bounded between 0 and 1. Additionally, it included a weak normal prior for the parameter $\eta$ with mean 0 and standard deviation 0.1.

Accordingly, for a modulation of the reward sensitivity, beta was adapted similarly, with the addition of the state value scaled by a run-invariant, subject-specific weight parameter $\eta$.

$$\beta'^{i} = \beta^i + \eta^s * \text{state}^i \qquad 3)$$

again with the boundary implemented ensuring that run-level beta estimates were positive. For the additional weight parameter $\eta$, a weak prior was included, drawn from the standard normal distribution.

To improve model fit, we explored multiple scalings of the state variables and the run number (e.g., rescaling to be between 0 and 1, z-scoring, z-scoring per ID and log transformation for the run-number).

***Model fit***.

The extended models were run with four chains and 5000 iterations each, including 2000 warmup iterations that were discarded instead of used for inference. As above, diagnostic values were checked, requiring no divergences to have occurred and all $\widehat{R} \leq 1.01$, ESS > 400, and BMFI > 0.2, along with diagnostic plots such as trace plots, energy plots and bivariate density plots. The extended model was again evaluated by comparing its results to the basic model fit.

For the learning rates, none of the models performed acceptably (with multiple variants of the scaling of state values as well as prior distributions). All of these models showed severely problematic behavior. Every attempt to fit one of these models resulted in many divergences, as well as large $\hat{R}$ values (around 4-5) and very small effective sample sizes, with estimated ESS around 2 for many parameters. To illustrate, for a direct extension of the basic model, with separate $\eta$ for win and loss learning rates and a weak normally distributed prior, fitted for 4000 iterations, there were 8000 post-warmup transitions, with a maximum $\hat{R}$ value of 4.51 and estimated ESS of parameter values ranging from 2 to 19.3.

For the reward sensitivity $\beta$ according to the participant's current metabolic state, weighted by the parameter $\eta$, performed better than the models modulating the learning rate. However, there were still more than 100 $\hat{R}$ value > 1.05 for parameters of interest (i.e., the individual $\eta$) for all potential modulators. Therefore, we decided to run post-hoc mixed-models in brms to estimate individual adaptations to mood, metabolic state, or over runs also including multiple predictors in one model.

### Figures

#### Non-negative matrix factorization reveals multidimensional structure of eating behavior related to obesity and binge eating

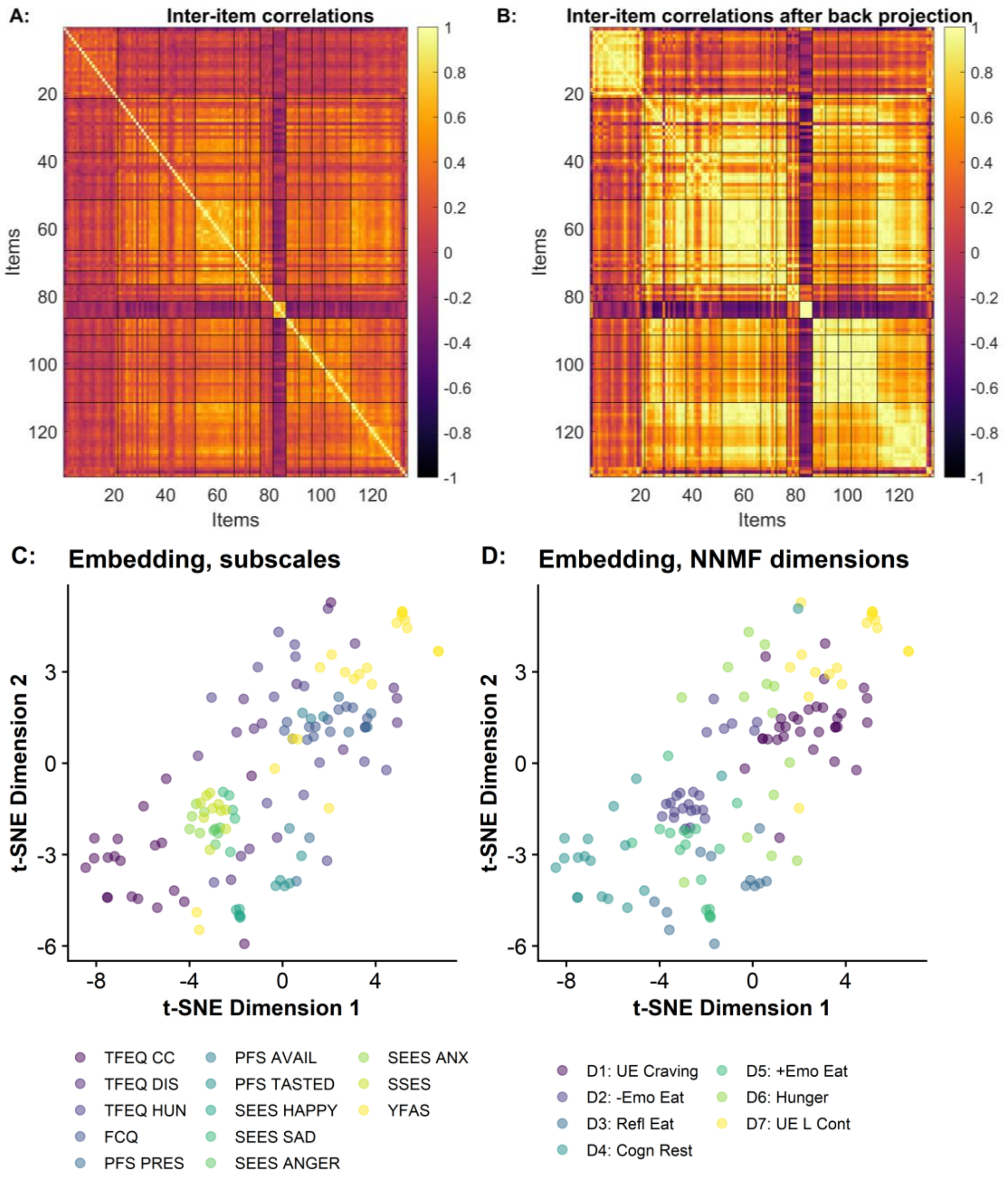


**Figure S1:** Non-negative matrix factorization (NNMF) captures the dimensionality of eating behavior effectively. A: Inter-item correlations for each item taken from the questionnaires TFEQ, FCQ, PFS, SEES, SSES, and YFAS. Items are ordered according to subscales and lines depict boundaries. B: Recovered inter-item correlations after multiplicative back-projection of the results of the dimensionality reduction via NNMF. The similarity of the inter-item correlation matrices indicates a good recovery of the latent dimensions across questionnaires. C: To visualize the item-level data in a low-dimensional space, we applied t-distributed stochastic neighbor embedding (t-SNE). Most items clustered close to other items of their subscale (color coded), but many items also showed a closer proximity to related subscales. D: Items assigned to the NNMF dimension with the highest loading (color coded) showed a well-aligned organization within the low-dimensional space that improved upon subscales.

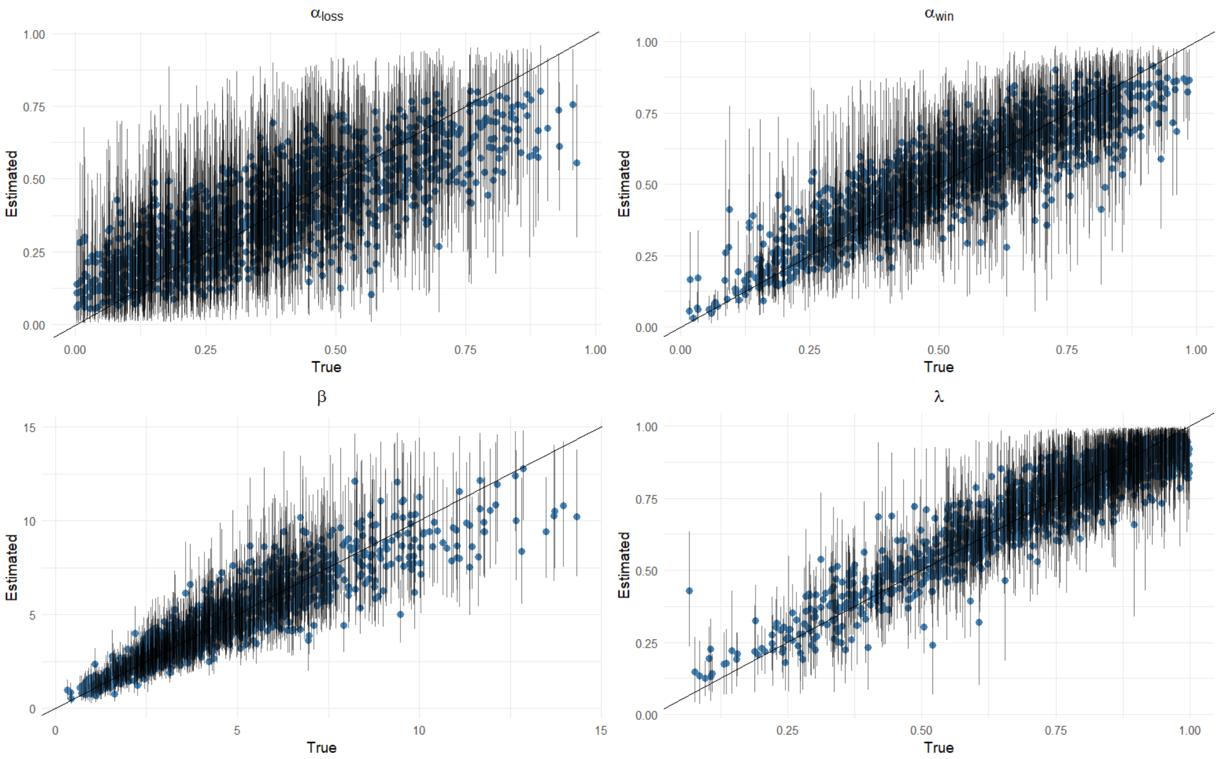


**Figure S2**: Parameter recovery plots for the four model parameters. Blue dots correspond to estimated (y-axis) and true (simulated) values (x-axis), while vertical lines show 90% equal-tailed Credible Intervals of the estimated parameters.

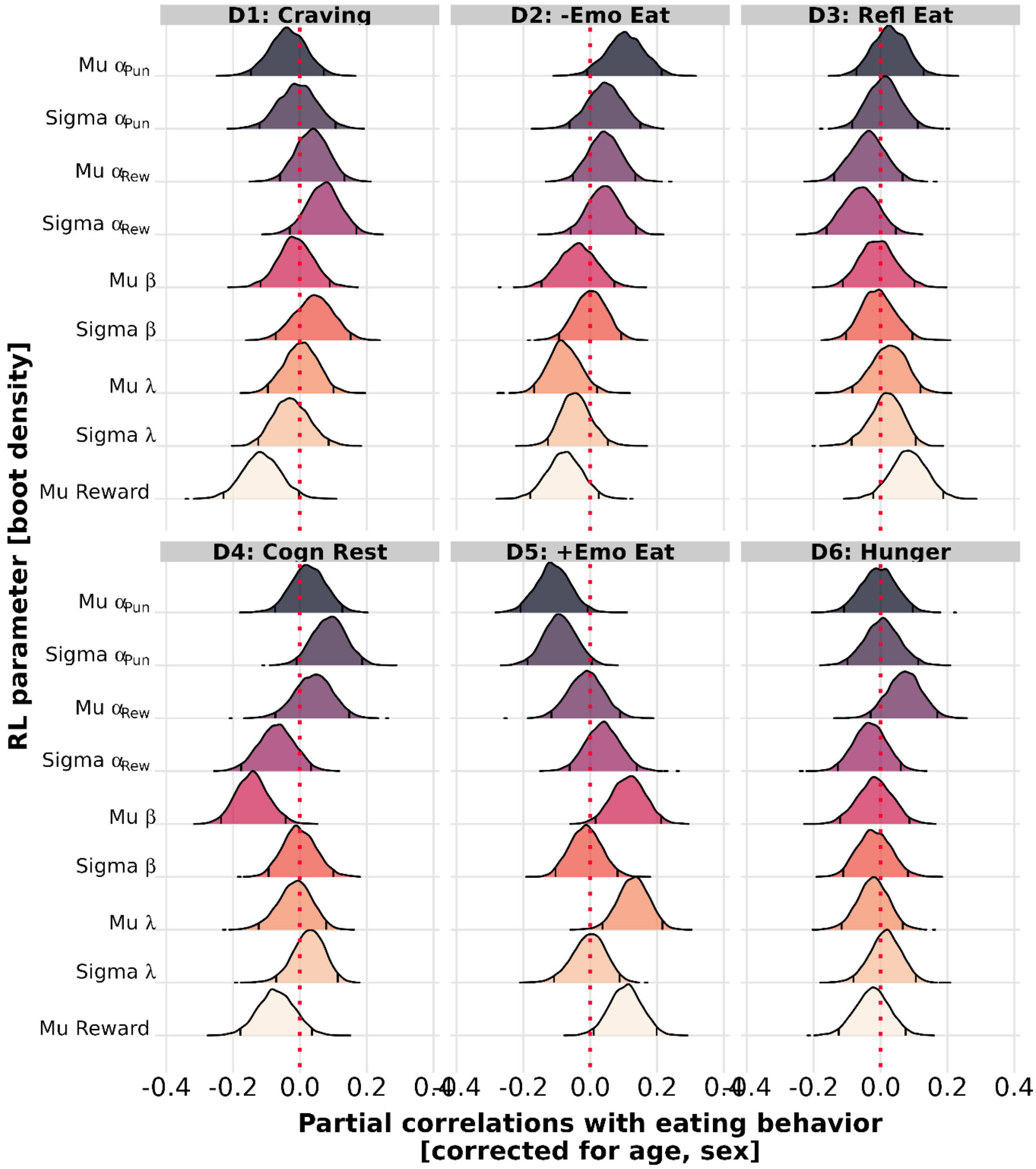


**Figure S3:** Bootstrapped partial correlations of the dimensions of eating behavior with reinforcement learning parameters corrected for age, and sex. Variability estimates are residualized for differences in the average of the parameter. Distributions show bootstrapped (5,000 resamples) distributions of the partial correlations with 2.5 and 97.5 percentiles.

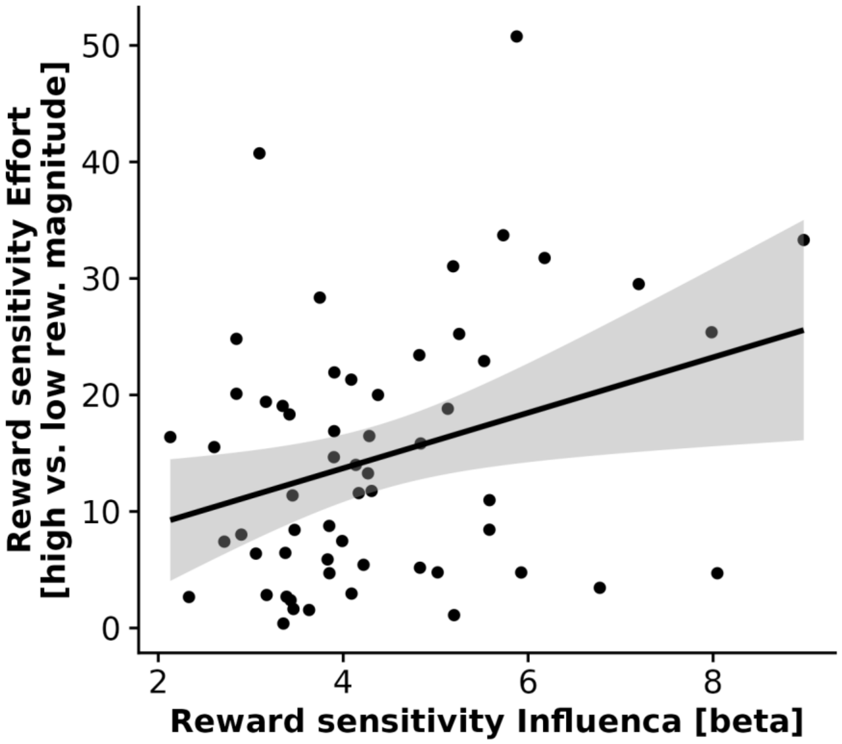


**Figure S4:** Average reward sensitivity, β, derived from the reinforcement learning task is associated with the adjustement of effort expenditure based on the reward magnitude at stake capturing reward sensitivity independent of learning (*r*(56)=.31, *p*=.018).

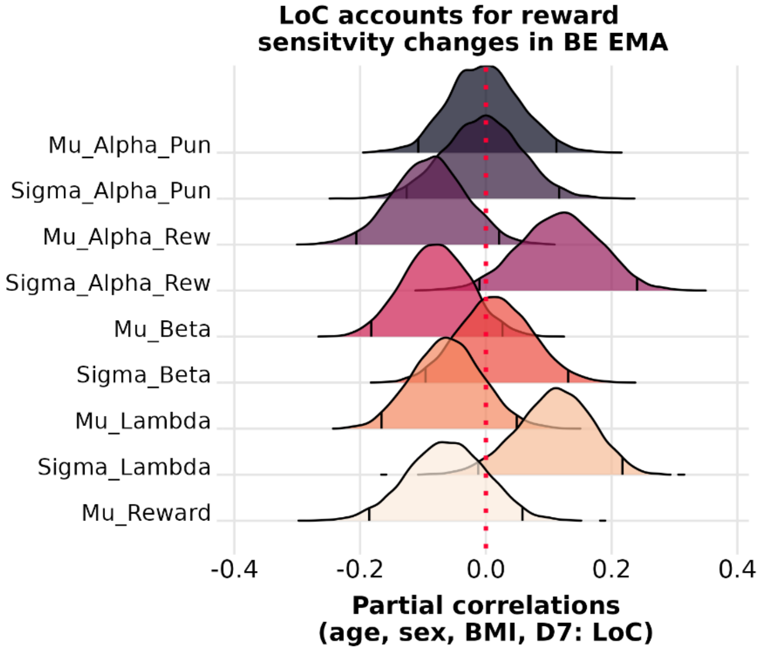


**Figure S5:** Accounting for trait-like positive loss of control reduces associations of binge eating during the online assessment (BE EMA) and reward sensitivity average and variability. Partial correlations corrected for age, sex, BMI, and loss of control show no significant associations of BE EMA with any of the learning parameters. Variability estimates are residualized for differences in the average of the parameter. Distributions show bootstrapped (5,000 resamples) distributions of the partial correlations with 2.5 and 97.5 percentiles.

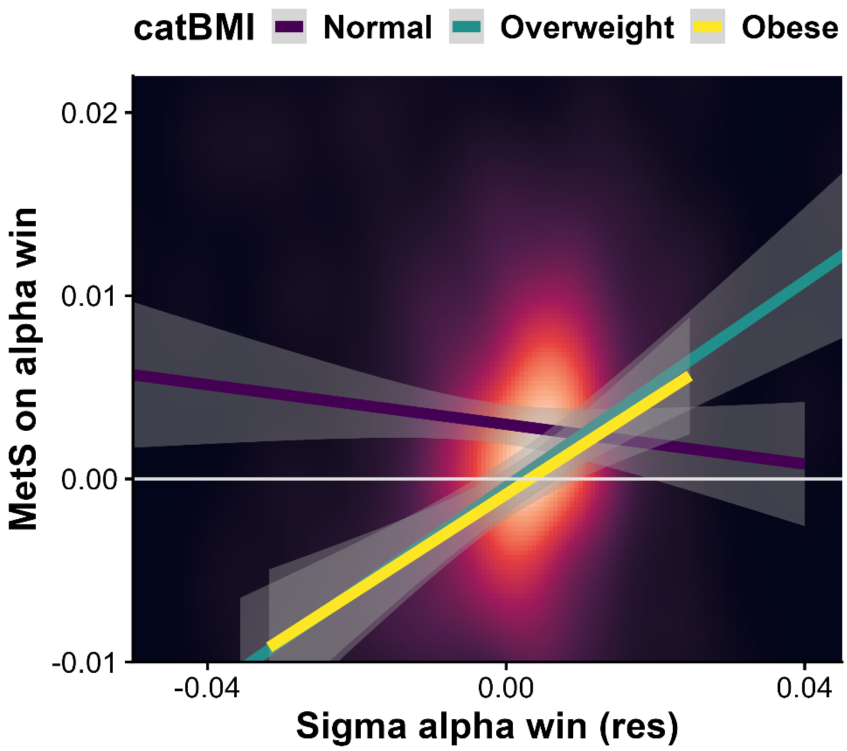


**Figure S6:** Variabilities in the learning rate for wins are associated with the metabolic scaling of the learning rate for wins in participants with overweight and obesity but not in normal-weight participants (BMI x SD: *b*=0.002, *p*<.001). The density plot shows the distribution of observations with lighter colors indicating higher densities. The shaded areas depict 95% confidence intervals.

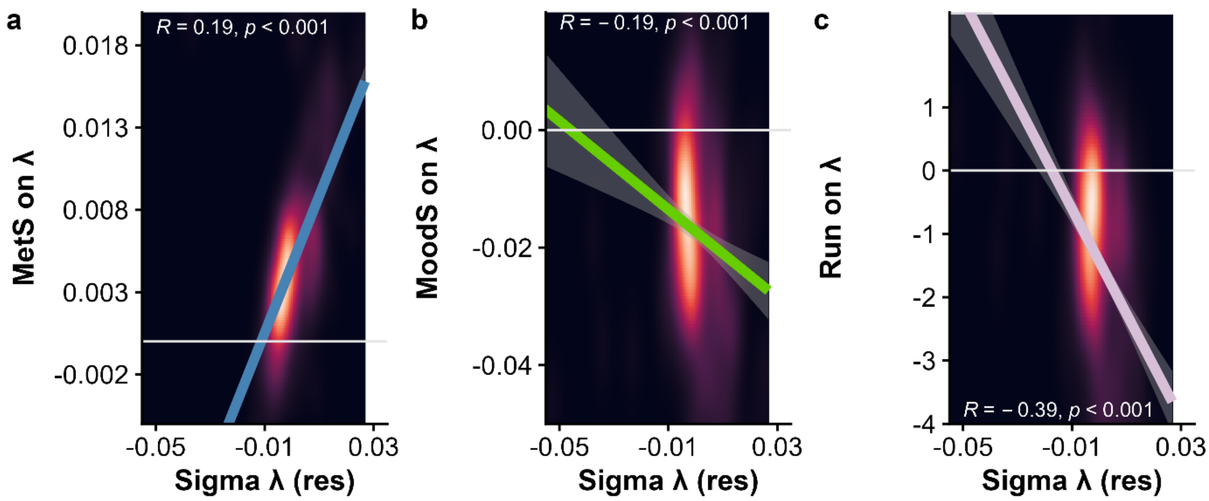


**Figure S7:** Variability in reinforcement learning (RL) lambda is partly explained by improvements over runs and state effects of mood and metabolism. Note that the run effect is estimated as an exponential decrease and thus negative values indicate larger increases over runs.

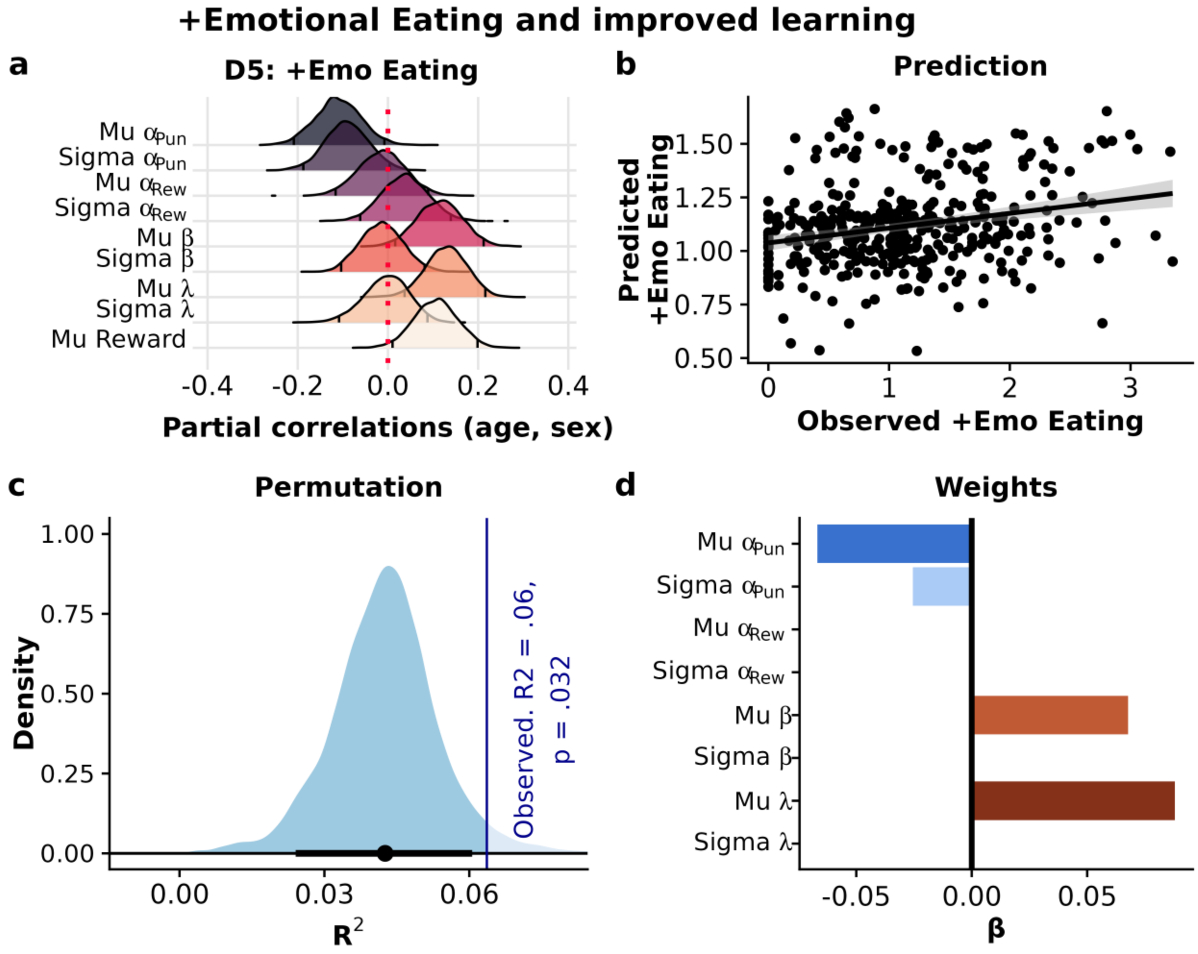


**Figure S8:** Trait-like positive emotional eating is associated with advantageous behavior in the task. a) Partial correlations corrected for age, and sex show significant associations of positive emotional eating with more points won, higher reward sensitivities, and higher reliance on learned values instead of reward magnitude (i.e., λ) as well as lower learning rates for punishments. Variability estimates are residualized for differences in the average of the parameter. Distributions show bootstrapped (5,000 resamples) distributions of the partial correlations with 2.5 and 97.5 percentiles. b) A model including learning parameters predicted positive emotional eating. Predicted and observed values of positive emotional eating were correlated and the model explained 6% of the total variance. c) Adding reinforcement learning parameters improves the prediction of positive emotional eating compared to the confounding variables age and sex. Error bars depict 95% percentiles. d) Feature weights contributing to the prediction of positive emotional eating. Higher reward sensitivity and λ together with lower learning rates for punishments contribute to the prediction.

### Tables

**Table S1**: Elastic Net model performance for all dimensions of eating behavior predicted by reinforcement learning parameters.

| **Dimension of eating behavior** | **$\Delta R^2$** | **$p_{perm}$** |
|---|---|---|
| Craving | .005 | .27 |
| -Emotional Eating | .0005 | .47 |
| Reflective Eating | .01 | .094 |
| Cognitive Restraint | .006 | .062 |
| +Emotional Eating | .02 | .033 |
| Hunger | .007 | .052 |
| Loss of control | .022 | .036 |
